# Systematic surveillance of SARS-CoV-2 reveals dynamics of variant mutagenesis and transmission in a large urban population

**DOI:** 10.1101/2024.07.25.24311007

**Authors:** Marie-Ming Aynaud, Lauren Caldwell, Khalid N. Al-Zahrani, Seda Barutcu, Kin Chan, Andreea Obersterescu, Abiodun A. Ogunjimi, Min Jin, Kathleen-Rose Zakoor, Shyam Patel, Ron Padilla, Mark Jen, Princess Mae Veniegas, Nursrin Dewsi, Filiam Yonathan, Lucy Zhang, Amelia Ayson-Fortunato, Analiza Aquino, Paul Krzyzanowski, Jared Simpson, John Bartlett, Ilinca Lungu, Bradly G. Wouters, James M. Rini, Michael Gekas, Susan Poutanen, Laurence Pelletier, Tony Mazzulli, Jeffrey L. Wrana

## Abstract

Highly mutable pathogens generate viral diversity that impacts virulence, transmissibility, treatment, and thwarts acquired immunity. We previously described C19-SPAR-Seq, a high-throughput, next-generation sequencing platform to detect SARS-CoV-2 that we deployed to systematically profile variant dynamics of SARS-CoV-2 for over 3 years in a large, North American urban environment (Toronto, Canada). Sequencing of the ACE2 receptor binding motif and polybasic furin cleavage site of Spike in over 70,000 patients revealed that population sweeps of canonical variants of concern (VOCs) occurred in repeating wavelets. Furthermore, we found that subvariants and putative quasi-species with alterations characteristic of future VOCs and/or predicted to be functionally important arose frequently, but always extinguished. Systematic screening of functionally relevant domains in pathogens could thus provide a powerful tool for monitoring spread and mutational trajectories, particularly those with zoonotic potential.

## Introduction

Repeated outbreaks of respiratory viral infections over the past 20 years culminated in the COVID-19 pandemic caused by SARS-CoV-2^1^, with more than 700 million cases and over 7 million deaths confirmed worldwide^2^. SARS-CoV-2 thus presented unique challenges for public health agencies trying to manage both travel and local spread. In particular, long incubation and recovery times coupled with large numbers of asymptomatic/mildly symptomatic patients that unknowingly communicate disease drove the pandemic to unprecedented levels.

The RNA SARS-CoV-2 virus is a member of the coronavirus family characterized by high mutational rates that lead to genetic diversity. The main variants impacting transmissibility, severity, and/or immunity are classified as Variants of Concern (VOCs). The first VOC, Alpha (B.1.1.7) was identified in September 2020 in the United Kingdom^3, 4^, followed by the identification of other VOCs that arose in a variety of regions including Beta (B.1.351) in South Africa in May 2020^5^, Gamma (P.1) in Brazil in November 2020^6^, Delta (B.1.617.2) in India in October 2020^2^, and Omicron (B.1.1.529) arising in various countries in November 2021^2^. The continuous process of genetic variation, competition, and selection results in the detection of major and minor subvariants within these VOCs^7,8^. The equilibrium-like status of a given VOC collapses when an advantageous new mutant emerges and outcompetes the pre-existing VOC within the population.

As the scale of SARS-CoV-2 transmission spiralled, the capacity of whole genome approaches to systematically map genetic alterations at a population scale was quickly overwhelmed, financially unsustainable, and slow, making it difficult to track emerging new variants until they had already escaped into broader regional and then global populations. We previously developed C19-SPAR-Seq (COVID-19 screening using Systematic Parallel Analysis of RNA coupled to Sequencing)^9^ as a high throughput next-generation sequencing (NGS)-based strategy to enable rapid detection of SARS-CoV-2. Therefore, as variants began emerging in the Fall of 2020, we pivoted the application of C19-SPAR-Seq to mapping the evolutionary dynamics of two key functional regions of SARS-CoV-2 in a large North American urban center. Here, we describe optimization and automation of the C19-SPAR-Seq pipeline and describe its use in providing near real-time profiling of 73,510 SARS-CoV-2-positive patients collected within the Greater Toronto Area (GTA; ∼6M people) from December 2020 to March 2023. This screening effort identified the early emergence of VOCs and revealed repeated acceleration and deceleration phases of VOC transmission that were unrelated to non-pharmaceutical interventions (NPIs). Furthermore, we show that subvariants frequently arise within VOC populations, but never spread, and that by tracking subvariants and minor putative viral quasispecies, alterations that foreshadowed future evolutionary SARS-CoV-2 trajectories could be revealed. Systematic profiling of functional domains in highly mutable viruses could thus provide an important tool to prevent and manage future pandemics.

## Results

### Integration of C19-SPAR-Seq into a Clinical Pipeline

We previously established a C19-SPAR-Seq platform for high throughput detection of SARS-CoV-2^9^ in which amplicons were designed over the RNA-dependent RNA polymerase (*RdRP*), and two key functional regions of the Spike (*S*) gene: the receptor-binding motif (*S-Rbm*) and the furin cleavage site (*S-Pbs,* FCS). In light of the global emergence of SARS-CoV-2 VOCs in the fall of 2020^5, 6, 10^, we set out to link C19-SPAR-Seq into a clinical diagnostics pipeline (Clinical Diagnostics Lab, Department of Microbiology at Mount Sinai Hospital and University Health Network) that provided COVID19 diagnostics services to the Greater Toronto Area (GTA) (**Fig. 1a, Supplementary Fig. 1**, **Supplementary Table 1**). Starting in December 2020 we profiled 1,218 SARS-CoV-2 positive samples collected between December 17, 2020, and January 28, 2021, then subsampled an average of 111 positive samples per week between January 28, 2021 and May 19, 2021, prior to transitioning to testing all positive samples in June 2021 (**Fig. 1a**; **Supplementary Fig. 1**). Importantly, we did not pre-select samples based on C_T_ values from the initial qRT-PCR tests, but established several quality control cutoffs: 1) total read counts for each amplicon within a given sample were required to be greater than that of the negative control within each plate; 2) read counts for *S-Rbm* had to be greater than two times the absolute deviation above the median counts for *S-Rbm* within each plate; and 3) samples with low viral-reads were filtered out if a sample’s total viral counts were lower than the total viral counts of the negative control within each plate. Of the initial 1,218 samples, only 20 failed to pass our quality control filters, representing less than 2% of the samples (**Supplementary Table 2**). Importantly, the remaining 98% of samples showed a high proportion of reads for all four amplicons (**Fig. 1b**). To call variants in C19-SPAR-Seq data, we analyzed the top read count for each amplicon following deep sequencing, and first focused on the major VOCs highlighted by the WHO^11^ that circulated in the global population in early 2021. This showed that most of the samples had *S-Rbm* and *S-Pbs* sequences from the original Wuhan strain (hereafter referred to as “wild type”, WT). However, we also identified the first cases of the WHO-designated variants of concern (VOCs), Alpha and Beta, detected in the GTA (**Fig. 1c,d**) which we confirmed by whole genome sequencing (WGS) (**Supplementary Table 3**). Together, this pilot analysis of the first 1,218 patients validated the C19-SPAR-Seq pipeline as an effective, scalable tool to accurately identify mutations within two key functional domains of SARS-CoV-2-positive patient samples. Of note, almost all VOCs that emerged between 2020 and 2023 were distinguishable based on sequence variations in the *S-Rbm* and *S-Pbs* regions targeted by C19-SPAR-Seq version 1 (V1, **Supplementary Fig. 1b**).

**Fig. 1.**
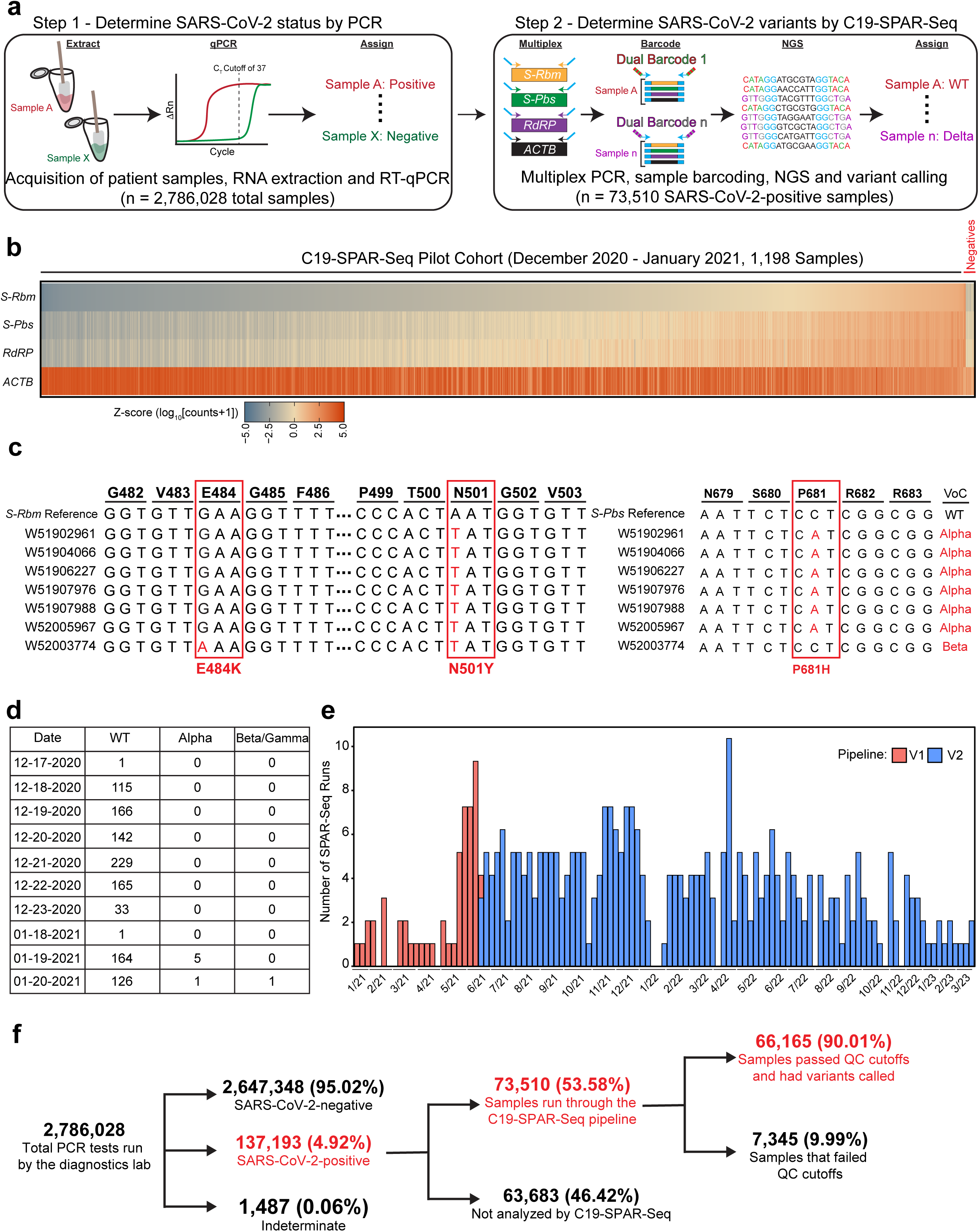
Integration of C19-SPAR-Seq to detect SARS-CoV-2 Variants of Concern. **a.** Schematic representation of the preclinical testing of patient samples for SARS-CoV-2 (Step 1) and the C19-SPAR-Seq multiplex sequencing pipeline (Step 2) to identify variants of concern. **b**. Analysis of SARS-CoV-2-positive patient samples by C19-SPAR-Seq V1. A pilot cohort (n = 1,198) was analyzed by C19-SPAR-Seq and the Z-score of log_10_ (read counts+1) for each of the indicated amplicons is presented in a heatmap. Samples are ordered from lowest to highest read counts for *S-Rbm*, with negative control samples on the right (n = 20). **c**. Representative alignment of top read counts for *S-Rbm*. Seven representative cases are shown aligned to the *S-Rbm* reference sequence, nucleotide substitutions are shown in red within the sequence and amino acid changes are highlighted in a red box. The Variant of Concern (VOC) that each sample contains is shown on the right of the sequence. **d**. Quantification of all major mutations observed in the pilot cohort. The frequency of each observed mutation is indicated. **e**. Number of C19-SPAR-Seq runs per week (Y-axis) from December 2020 to March 2023 (X-axis). Red bars are runs with the multiplex primer V1 and blue bars are runs with the multiplex V2 primers of the C19-SPAR-seq pipeline. **f**. Description of the number of samples run through the C19-SPAR-Seq (red).

### Optimization of an improved C19-SPAR-Seq V2 pipeline

Having benchmarked the C19-SPAR-Seq V1 pipeline, and to facilitate its integration into a clinical diagnostics platform, we next developed a semi-automated pipeline for the rapid, systematic screening of SARS-CoV-2 variants (**Supplementary Fig. 2**) that included semi-automated library generation, sequence processing, and variant assignment. Briefly, PCR-positive samples were arrayed in 384-well formatted plates, processed, sequenced, analyzed, and a report generated with the pipeline providing the capacity for variant profiling within 24h (**Supplementary Fig. 2**). This pipeline was applied to profile an average of 111 cases per week in early 2021 and was accelerated on June 1, 2021 to include all positive samples and daily screening by the summer of 2021, as increasing numbers of VOCs were being detected globally, and the more aggressive Delta variant had emerged (**Fig. 1e**). During the course of variant monitoring from December 17, 2020, to March 27, 2023, a total of 2,786,028 PCR tests were performed of which 133,089 (4.88%) were SARS-CoV-2-positive, and less than 0.05% of tests were indeterminate (**Fig. 1f**). Over the course of our analysis, we ran 73,510 of the 137,193 SARS-CoV-2-positive samples through the C19-SPAR-Seq platform; this represents 53.58% of all positive cases, with 81.48% profiled after June 1, 2021 (**Fig. 1f**). Importantly, we found that only 7,345 samples failed QC assessment despite not filtering for viral load (*ie*-Ct values). This represented a failure rate of ∼10% over the test period, showing that C19-SPAR-Seq can provide robust coverage of variants in a clinical diagnostic setting (**Supplementary Table 2**).

Over the course of screening we also tracked the analytical performance of our pipeline on a run-to-run basis and adjusted parameters to improve our standard operating procedures as necessary (see Methods). For example, while the *S-Rbm* and *S-Pbs* regions produced fingerprints that covered most VOCs, including most Omicron family members (see below), this was not the case for Delta. Although Delta harbours a P681R alteration in the *S-Pbs* that was captured, the *S-Rbm* region originally screened was WT, and the characteristic T478K mutation in the *S-Rbm* was not covered (**Supplementary Fig. 1b**). When Delta emerged as a global threat, we rationalized that our pipeline was amenable to changes in primer sequences that would capture new regions of interest, and designed a new primer pair (*S-Rbm* V2 and pipeline version V2) which yields a larger amplicon (**Supplementary Fig. 3a**) that covers an additional five amino acids (including T478) that contribute to ACE2 receptor binding^12, 13^. We validated that the *S-Rbm* V2 primer pair performed well in our C19-SPAR-Seq assay by qRT-PCR (**Supplementary Fig. 3a**) and multiplexed amplicon distribution by fragment analyzer (**Supplementary Fig. 3b; right panel**). One limitation of the V1 primer mixture was the high level of non-specific amplification that was observed in the multiplex primer reaction (**Supplementary Fig. 3b**; left panel), which required a size selection purification step using the Pippin Prep System to purify the 220-350 bp fragment window prior to sequencing. However, the V2 primer mixture that increased the *S-Rbm* amplicon by 35 bp, provided more specific amplification products from the multiplex reaction (**Supplementary Fig. 3b; right panel**), obviating the need for a size selection step. Systematic mapping of primer combinations could thus improve technical performance in future iterations of C19-SPAR-Seq. Next, we validated V2 against V1 in a diagnostic setting in four clinical runs, which contained a total of 94 samples. Comparison of both V1 and V2 in our C19-SPAR-Seq pipeline showed good correlation of *S-Rbm* reads (Spearman Correlation of 0.82, r^2^=0.67; **Supplementary Fig. 3c**) without affecting the efficiency of the other primer sets (**Supplementary Fig. 4**). Most importantly, mutation and variant calling was the same for each of the samples, but with the added benefit that V2 provided direct sequence information around the T478K mutation found in Delta (**Supplementary Data 2**).

Another potentially confounding factor in large-scale sequencing experiments is the risk of sample mixing and/or crosswell contamination during the experimental setup and workflow. To directly measure cross-well contamination rates in the PCR step we developed *S-Rbm* V2 primer pairs that incorporated well-specific molecular identifiers (barcodes; BC-*S-Rbm* V2) (**Supplementary Fig. 5a, Supplementary Table 4**). The 384 BC-*S-Rbm* V2 primer pairs were validated by qPCR on a control sample (data not shown) and were employed to re-analyze 6 clinical runs (1,987 positive samples) across the testing window that contained a representative mixture of WT, Alpha, Delta, and Omicron B.1.1.529 VOCs. Of note, BC-*S-Rbm* V2 primers did not negatively affect library quality (**Supplementary Fig. 5b**). We then measured contamination in each well as the percentage of reads that do not contain the correct barcode pair. Across all these barcoded clinical runs, we observed that 97% of all reads belonged to the correct well-barcode pair (**Supplementary Fig. 5c, Supplementary Data 1**). Importantly, samples which contain a low percentage of correct well-barcode pairs all contained extremely low total read counts, indicative of a failed PCR reaction. Therefore, these samples would later be filtered out for having poor quality in the analysis pipeline (**Supplementary Fig 6**). In addition, we measured how often paired well-barcodes within a plate were found in the incorrect well. This revealed that on average 2.5% of the total reads for any given sample could be attributed to cross-well contamination (**Supplementary Fig. 5d**). Importantly, mapping the spread of well-barcode mixing events revealed a stochastic pattern of contamination across six independent clinical runs, indicating that there is not a systematic error within our robotics pipeline and sample handling (**Supplementary Fig. 7**). Together, these studies show that the C19-SPAR-Seq pipeline is highly sensitive, accurate, robust and adaptable to rapid modifications in the primer multiplexing step, which is extremely advantageous when characterizing highly infectious and mutable viruses such as C19-SARS-CoV-2.

### Population-level surveillance of variant dynamics by C19-SPAR-Seq

To track variant spread, we next integrated sequence information into a single dataset (See methods). Each major VOC was detected within our jurisdiction, albeit with different latencies from the first emergence worldwide (**Supplementary Table 5**). Of the VOCs identified in the GTA, only Alpha, Delta, and four Omicron sublineages generated dominant waves (**Fig. 2, upper panel**). Of the dominant VOCs, we noted that only a single variant was dominant at any time, as noted in other jurisdictions (**Fig. 2, lower panel**)^14, 15^. We next assessed the transition between variants by calculating the cross-over point when each subsequent dominant VOC became proportionally more of the tracked cases than the previous dominant VOC (**Fig. 2, lower panel**).

**Fig. 2.**
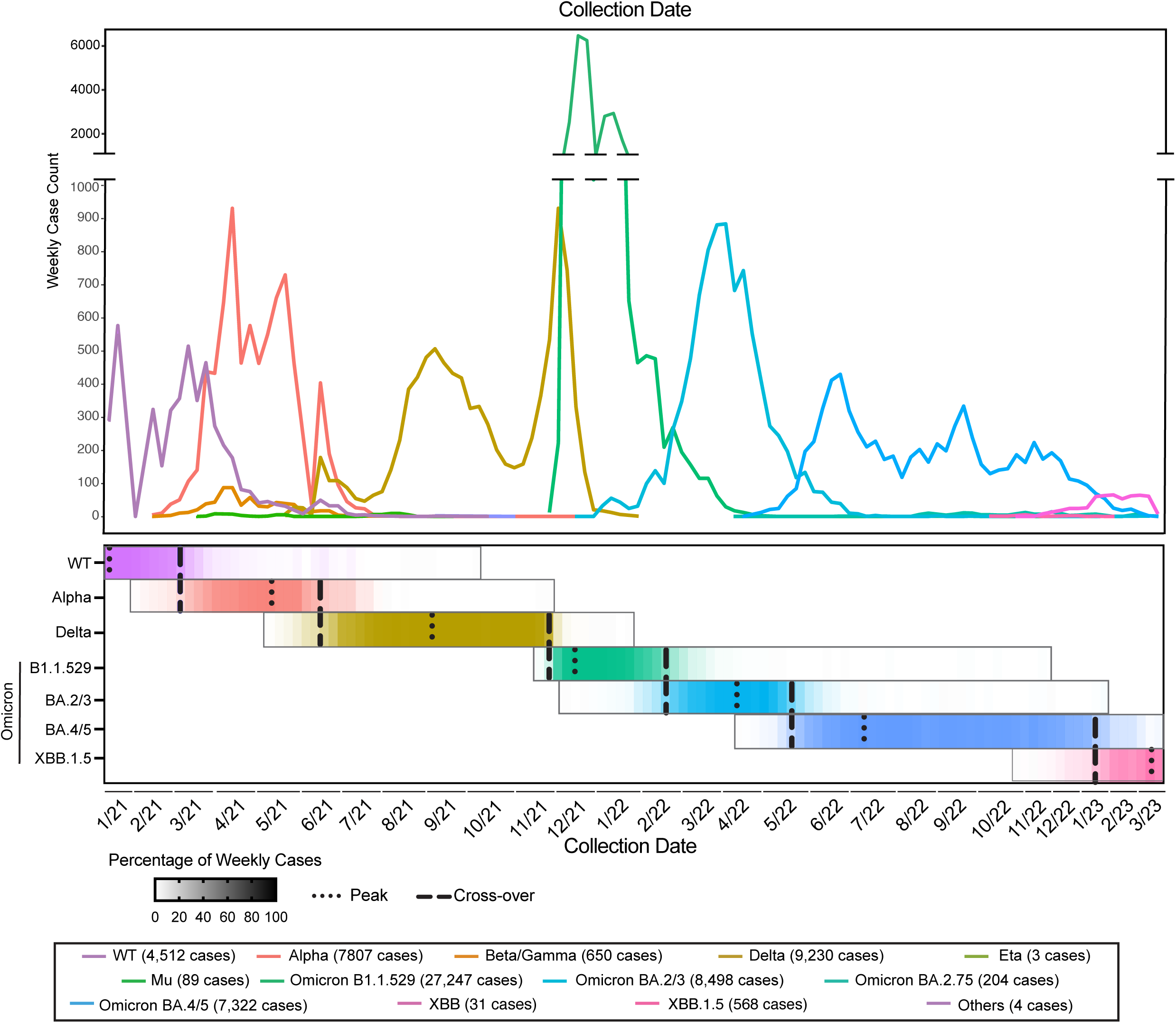
Results of an Improved C19-SPAR-Seq V2 pipeline. Line graph of the total number of cases for each canonical VOC from December 2020 to March 2023 (upper panel). Percentage of the weekly cases of the variants, the peaks and cross-over are indicated with dot and dashed lines respectively (lower panel).

This showed a shift from WT to Alpha in late February 2021, from Alpha to Delta in June 2021, and Delta to Omicron B.1.1.529 in December 2021. As expected, Omicron numbers and frequency dominated the population of variants by late December 2021, with subsequent Omicron subvariants emerging with their first reported cases on December 23, 2021 (BA.2/3), May 6, 2022 (BA.4/5), November 16, 2022 (XBB), and December 8, 2022 (XBB.1.5) (**Fig. 2, upper panel, Supplementary Table 5**). In contrast to these major VOCs, the Beta and Gamma variants displayed low steady-state infectivity of <10% in the Spring of 2021, before extinguishing on August 7, 2021, and we grouped them together for our analyses (**Fig. 2**). Furthermore, Mu (B.1.621.1), which arose in Columbia in January 2021 was only detected in 89 samples, peaking in August 2021, while Eta (B.1.525) which arose in the United Kingdom and Nigeria in December 2020 was only detected in 3 samples (**Fig. 2, upper panel**). Given their low frequency these latter two were not further characterized in this study.

We postulated that the unique sequences of variants, combined with systematic, high frequency screening, would provide a sensitive method to track the dynamics of SARS-CoV-2 spread within the population. To examine this, we first focused on Alpha, the first major VOC that circulated in the GTA and tracked its emergence and expansion based on the proportion of daily cases from February 6, 2021 (Day 1), until Alpha became the dominant species (>50%; Day 32; March 10, 2021; **Fig. 3a**). We next fit linear models along a sliding 5-day window (**Fig. 3a, top panel, red lines**) to quantify the expansion rate (Fig. 3a, bottom panel, see methods for details). This showed expansion rates were low in the beginning indicating a lag-phase in variant expansion. Interestingly, we observed that Alpha did not expand at a homogeneous rate within the population, but displayed distinct waves of accelerating and decelerating transmission (**Fig. 3a, lower panel**). Analysis of the emergence of Delta (**Fig. 3b**) similarly showed highly dynamic behaviour and acceleration/deceleration waves analogous to Alpha dynamics (**Fig. 3b, lower panel**). This was followed by the emergence of Omicron, which displayed a lag and then two phases of rapid acceleration (**Fig. 3c**). These data show that viral spread is comprised of multiple, repeated wavelets of transmission.

**Fig. 3.**
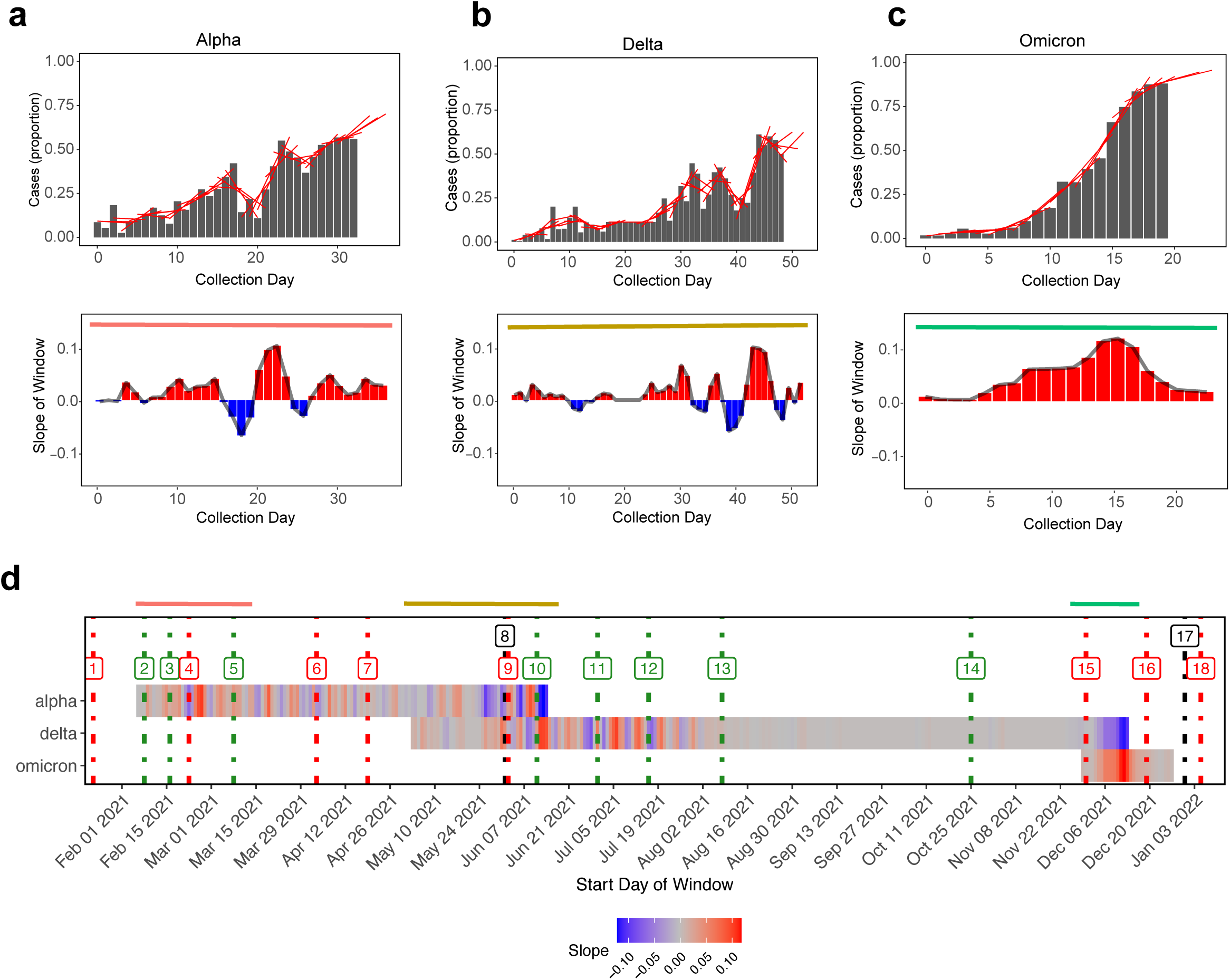
Variant dynamic and Non-Pharmaceutical Interventions (NPIs). **a.** Schematic of the procedure to track the dynamic spread of the virus (see Methods) showing the proportion of Alpha, Delta and Omicron cases with representation of linear models along sliding 5 days windows (red line) (upper panel). Slopes from each linear model (lower panel). **b**. Dynamic spread using slopes of WT, Alpha, Delta and Omicron B1.1.529. NPIs are plotted (red lines indicate closures or restrictions and green lines indicate openings; see **Supplementary Table 6**)

The distinct patterns of viral spread prompted us to explore whether acceleration waves were linked to NPIs. We therefore assessed major NPI policies implemented in the GTA in 2021, in particular, stay-at-home orders and school openings and closings (**Supplementary Table 6**). We overlaid NPI dates on our rate of change data for WT and the three major VOCs (**Fig. 3d**) and found that changes in NPI policies were not associated with alterations in the patterns of viral expansion. For example, between a closing NPI (#4; return to stay-at-home order) and an opening NPI (#5; stay-at-home order lifted), we noted 2 acceleration waves of Alpha, while the opening NPI (#5) was followed by a mixed period of reduced rates of transmission. Similarly, Delta cases were initially dynamic until July 2021, but transitioned to a steady-state rate of transmission despite societal-wide reopening that included the lifting of restrictions on gatherings (#13 to #14, August to September 2021; **Fig 3d, Supplementary Table 6**). Finally, the rapid onset of Omicron, as initially observed (**Fig. 2 upper panel**) showed an extreme acceleration and attempts to dampen Omicron spread by restricting travel (#15, **Fig. 3d**) were followed by a rapid acceleration phase. Indeed, systematic, rapid C19-SPAR-Seq screening showed substantive numbers of Omicron were already present in the GTA. These analyses indicate that patterns of viral spread are not strongly affected by NPIs and further suggest how near real-time monitoring of viral species can provide an important tool to inform decisions around the introduction of public health measure policies.

In December 2021, changes in provincial government policy that took effect on December 31, 2021 (#17, **Fig. 3d**), limited testing to high risk groups such as patients and staff in health care settings, or people living in or working in First Nation, Inuit, and Métis communities^16^, and significantly reduced population testing. Thus, subsequent frequencies may not reflect alterations in the general GTA population. This reduction in testing accessibility also had an immediate impact on our data collection. The total samples collected and sequenced between December 25 and December 31, 2021 was 6,813, while in the week immediately after testing restrictions (January 1 to January 7, 2022), only 1,035 samples were collected and processed, marking an 84% drop in samples. Due to the reduction in population-level data collection, we did not assess the impact of NPIs implemented after December 2021.

### Subvariants arise frequently in VOC populations

In the course of profiling known variant fingerprints, we noted that subvariants often arose with additional changes to the *S-Rbm* and/or *S-Pbs* sequences. For example, our preliminary screen of the first 1,219 samples revealed 3 VOCs with WT comprising 1,142 of the samples, Alpha (B.1.1.7) comprising 6, and Beta (B.1.351) comprising 1. Within this group we noted 46 WT samples that possessed 12 distinct patterns of alterations in the *S-Rbm* and *S-Pbs* (**Fig. 4a**). Three were mutations in *S-Rbm* (E484K, Y489Y, and S494P), eight had mutations in the *S-Pbs* (N679K, P681H, P681H/S691S, P681R, R682R, A684V, S691S, and I692I), and one had mutations in both regions (E484K/Q677H) (**Fig. 4a**). In contrast, analysis of the *RdRP* region that is also incorporated into the C19-SPAR-Seq amplicon set revealed 3 samples each with a unique variant sequence (**Fig. 4a**). To validate the C19-SPAR-Seq results in the pilot cohort, we confirmed the presence of these mutations by WGS (**Supplementary Table 3**), which showed these early variants all comprised the B strain and further showed that samples harbouring N679K alterations were closely related (**Fig. 4b**), suggesting they represent a small transmission cluster. In contrast, the other variants were more distant and may represent sporadic introductions and/or *de novo* emergence of alterations.

**Fig. 4.**
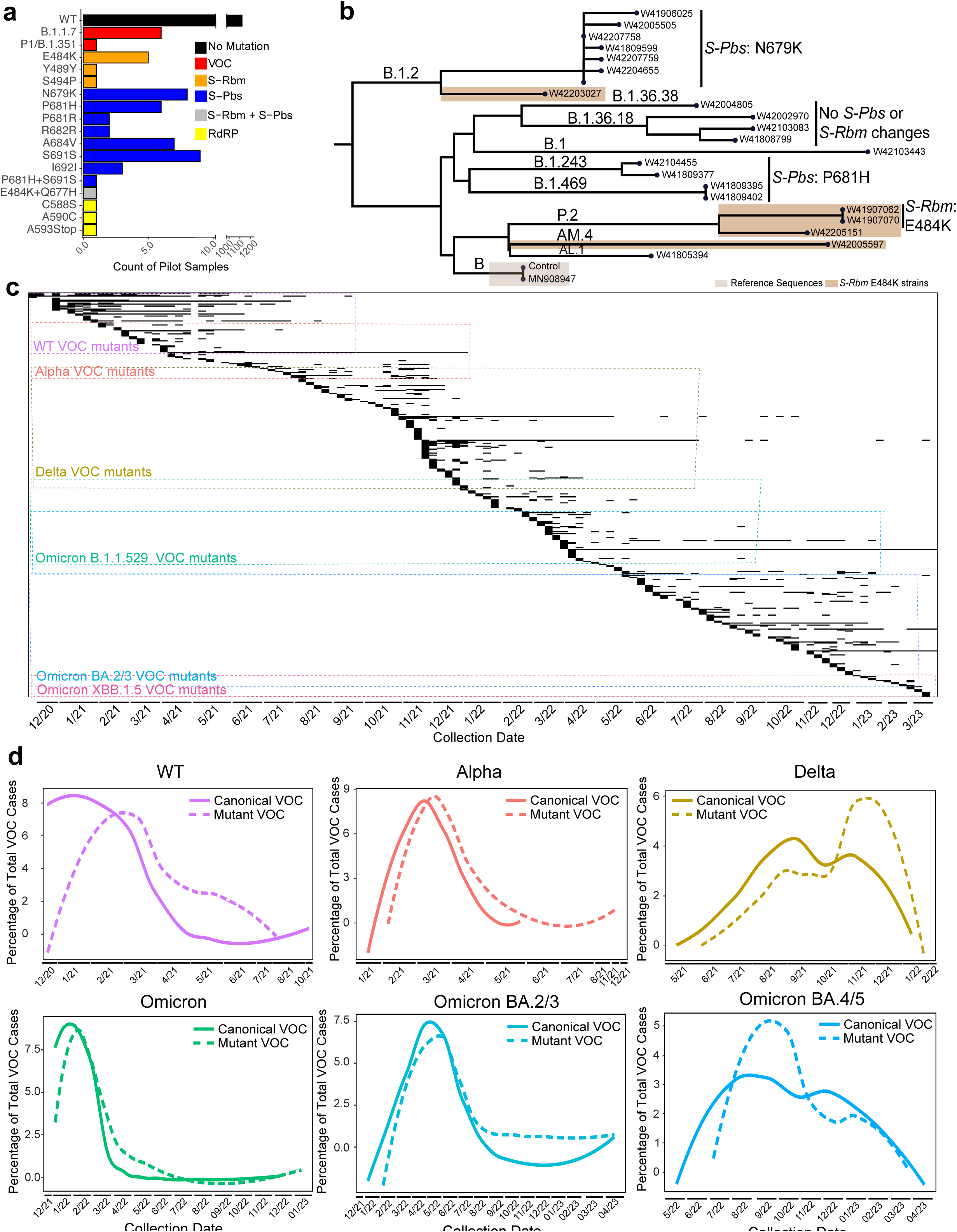
Population-level surveillance of variant dynamics by C19-SPAR-Seq. **a.** Quantitation of all major mutations observed in the pilot cohort. The frequency of each observed mutation is plotted and color-coded based on their location in the SARS-CoV-2 genome **b.** Phylogenetic tree of the WT strain and the early variants. **c.** Binary heat map of each canonical VOC and non-canonical VOC ordered by date of first detection. Within each coloured box is each mutant that shares a common VOC sequence trace. **d.** For each variant, the percentage of canonical VOC is shown with a solid line, and the percentage of mutant VOC is represented with a dash-line.

We next expanded our analysis and annotated the *S-Rbm* and *S-Pbs* of all SARS-CoV-2 isolates profiled in the GTA from January 2021-March 2023 (66,165 QC-passing samples). The WT and canonical VOCs accounted for 96.09% of all samples, with the remaining 3.91% of samples containing additional changes in the *S-Rbm* and/or *S-Pbs* (**Supplementary Fig. 8, 9, Supplementary Data 3**). In total we identified 393 non-canonical, unique alterations in VOCs, the vast majority of which (389 out of the 393 identified) were point mutations that changed the amino acid sequence (**Figure 4c, Supplementary Fig. 8, Supplementary Fig. 9d, Supplementary Data 3**), while the remaining 4 subvariants were an insertion and deletion in the *S-Pbs* (**Supplementary Fig. 10a**) and 2 silent mutations in BA.4/5 and BA.2.75.2 respectively. To identify these viral species and their dynamics within the population we next plotted each unique sequence throughout the time course (**Fig. 4c**) and referred to these as VOC subvariants **(Fig. 4c, Supplementary Fig. 8, Supplementary Data 3**). To assess if these mutational calls reflected poor-quality samples, we quantified the top read count percentages, which showed similar abundance for both canonical VOC and VOC subvariants across the entire dataset (Supplementary Fig. 10b). We also analyzed *RdRP* and only found 38 nucleotide variants, which formed 34 unique amino acid variants distributed in 194 samples (0.29%; **Supplementary Data 3**). This indicates that, unlike the *RdRP*, the *S-Rbm* and *S-Pbs* regions are subject to much greater mutational dynamics (Supplementary Fig. 9d), an observation consistent with evolutionary pressure converging on these key functional regions for SARS-CoV-2 transmission and virulence, and the fact that they are also primary targets of the immune system. In contrast to point mutants, we only identified five unique stop-gain mutations, which would be predicted to be disadvantageous for viral expansion (**Supplementary Data 3**). Similarly, while point mutations in the *S-Pbs* were frequently observed, we identified only 1 example of an insertion (seen in three samples), and one example of a deletion seen in two samples (**Supplementary Fig. 10a**). Thus, deletions and insertions within the *S-Rbm* and *S-Pbs* are extremely rare (**Supplementary Data 3**).

The dynamics of canonical VOCs within our data set shares the same patterns as those worldwide, where each subsequent VOC outcompetes the previous one (**Fig. 2**). However, we were curious as to how VOC subvariants behaved over time within the population. To determine this, we aggregated all subvariants for each unique VOC and plotted their frequencies within the population over time with respect to the canonical VOC. Interestingly, VOC subvariant frequencies peaked weeks after the peak of the canonical parental counterpart (**Fig. 4d**), suggesting that these subvariants arose within the population after the introduction of the canonical version. Interestingly, Delta and Omicron, which were two of the most dominant VOCs worldwide, had subvariants that formed secondary waves that peaked while the canonical variant was declining (**Fig. 4d**).

We next sought to map how specific mutations and/or residues within the *S-Rbm* or *S-Pbs* were being selected for, or recurrently altered over time. To investigate this, we calculated the frequency of mutations at each residue within the *S-Rbm* and *S-Pbs* amplicons for each VOC (**Fig. 5a**). One of the most striking observations was the high level of mutagenicity of the WT strain at positions E484, N501, N679, and P681, which together accounted for 79.3% of the WT VOC subvariants (**Fig. 5b, Supplementary Fig. 9b,c**). All of these residues were targeted for mutation in future VOCs, suggesting the WT virus was frequently generating variants in these functionally important residues. Furthermore, analysis of subvariant frequency showed that all VOCs frequently produced mutants, except the Beta/Gamma and XBB VOCs which displayed low levels of subvariant production (**Supplementary Fig. 9a,b**). Interestingly, Delta, which generated the most subvariants (**Supplementary Fig. 9b**), showed a high enrichment for mutations in the Omicron hotspots Q477, N501, and N679 (**Fig. 5a** and **Supplementary Data 3**). Omicron BA.2.75 also showed high production of subvariants (**Supplementary Fig. 9c**); the Omicron sublineage BA.4/5 often targeted Y473, Y489, A672, R683, and S691, most of which were readily seen in previous VOC subvariants, and may represent avenues for further evolution of SARS-CoV-2 (**Fig. 5b**). In contrast to *S-Pbs* and *S-Rbm*, we detected very few subvariants in *RdRP* (**Supplementary Fig. 9d**).

**Fig. 5.**
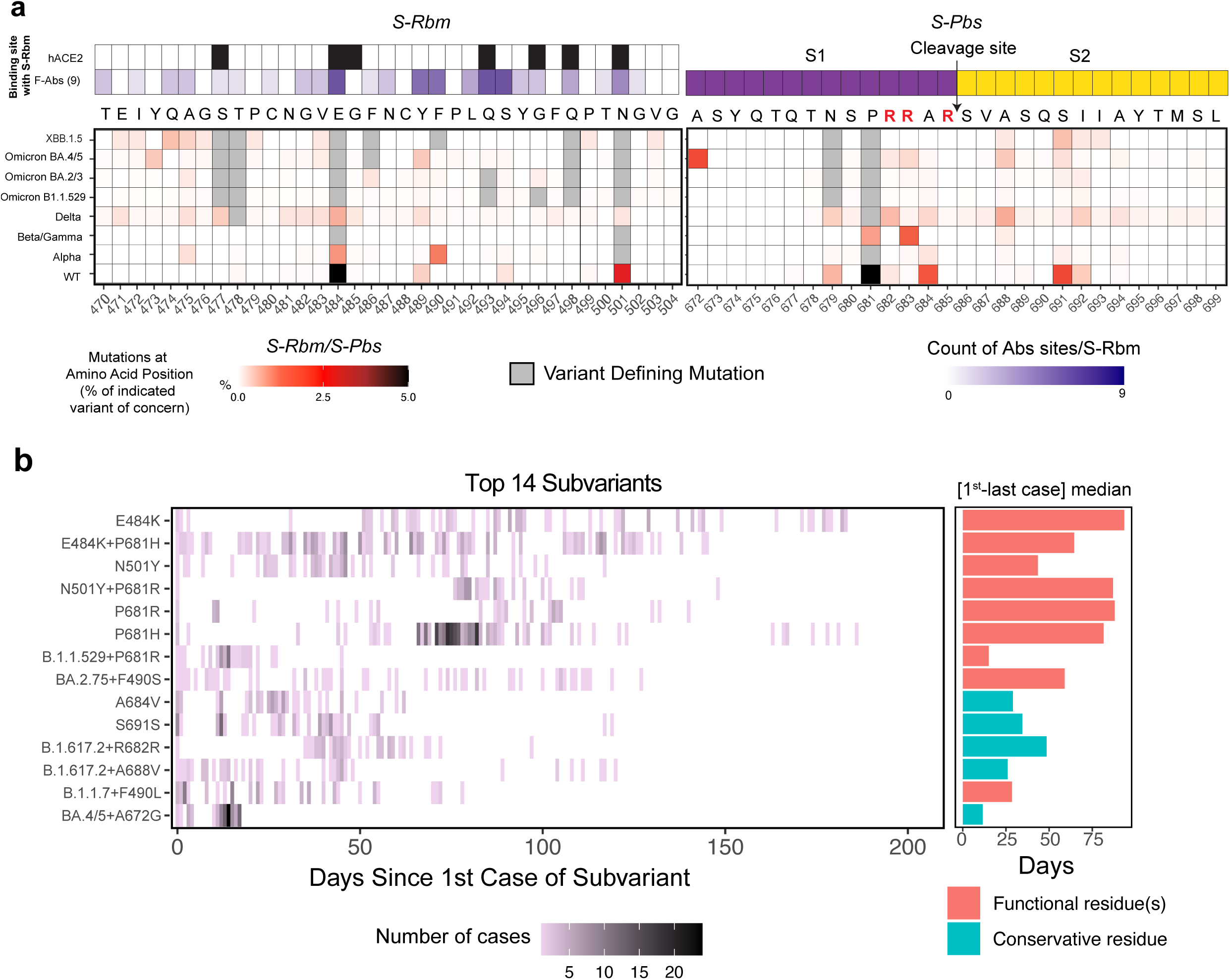
Major VOC subvariants within the *S-Rbm* and *S-Pbs* sequences. **a.** Percentage of top reads with a mutation detected in a given amino acid in *S-Rbm* and *S-Pbs*. The percentage of a point mutation at each amino acid position is shown. Canonical VOCs mutations are indicated in grey. The binding site of *S-Rbm* with hACE2 (black tiles) and 9 neutralizing antibodies are shown (white-purple scale, with darker purple indicating contact with more antibodies)^32, 33, 34^, as well as the *S-Pbs* cleavage site (S1 subunit indicated in purple, S2 subunit indicated in yellow) **b.** Heatmap of case counts of the top 14 key subvariants and the circulation time (time from the 1st-last case) are plotted with functional subvariants shown in red and non-functional subvariants shown in light blue.

To examine subvariant transmission within the population, we assessed the circulation time of the top 14 subvariants, which included both functional and non-functional mutations (**Figure 5b, left panel**). The median circulation time of subvariants with putative functional mutations (E484, P681, N501, F490) was 56 days compared to the non-functional group (A684, S691, R682, A688, A672) that displayed a median time of 29 days. This indicates that functional subvariants tend to circulate longer than their non-functional counterparts (**Figure 5b, right panel**). These observations suggest that alterations in specific residues recurrently arise *de novo* in the viral population, but without selection pressure, subsequently subside. To directly test this, we compared the affinity of ACE2 for S-RBMs corresponding to major VOCs, and a prominent Alpha subvariant we identified, Alpha + F490L. All of the VOCs except for Delta showed a higher affinity to ACE2 that reflected a slower off-rate for ACE2 binding *in vitro* **(Supplementary Fig. 11**). Furthermore, Alpha+F490L displayed similar binding kinetics as canonical Alpha, consistent with this variant *S-Rbm* not possessing a selective advantage. Rapidly identifying novel subvariants and assessing their functional impact can thus provide a useful tool to predict the dynamics of spread through the population. Collectively, these studies show that population-level surveillance of mutable viral pathogens, such as SARS-CoV-2 provides an extremely useful tool in predicting hotspot and key residues that will arise as major species in future lineages. Applying this strategy could thus provide important predictive value in monitoring viral pathogens with potential cross-over capability.

### Detection of putative SARS-CoV-2 quasispecies by C19-SPAR-Seq

The concept of quasispecies reflects a model of evolution in which imperfect replication leads to the production of mutant clouds that in aggregate define the evolutionary unit of selection^17^. In the context of viral evolution, a quasispecies represents a large collection of mutant genomes, with the dominant sequence reflecting the variant with the highest fitness within the cloud. As such, changes in selection pressure can alter which quasispecies dominate the population. C19-SPAR-Seq provides deep profiling of SARS-CoV-2 sequences over functionally important regions of the *S* gene and we assigned variants based on the major sequence (top read in the deep sequencing pipeline) obtained from each sample. These top reads accounted for approximately 75% of all mapped reads per sample on average across the entire dataset (**Supplementary Fig. 10b**), suggesting a significant number of minor subvariants were circulating in the population. Furthermore, using well-barcodes, we showed that the 25% of non-top reads were correctly assigned to the well, and thus do not represent crosswell contamination (**Supplementary Fig. 5c, d**). Since the quasispecies hypothesis posits that RNA-virus populations contain a high level of genetic variants that form a heterogeneous viral pool within a patient^18, 19, 20, 21^, we investigated these minor reads in greater detail. The abundance of these minor species is reported to be approximately 1%^22^, which is quantifiable using C19-SPAR-Seq, given the per-patient read depths that were on average 115,685 reads.

To search for putative quasispecies (pQS), we took advantage of the barcoded sequencing data we generated for six clinical runs that contained representative samples from WT, Alpha, Delta, and Omicron B.1.1.529 VOCs (**Supplementary Fig. 5c,d**) and selected 1,110 samples to perform minor sequence analysis on barcoded *S-Rbm* sequences as well as non-barcoded *S-Pbs* sequences. The *S-Rbm* barcoding allowed us to account for cross-contamination, which could contribute significant background noise that would confound identification of pQS. We established a run-specific multi-step filtering process to identify pQS within each sample (**Fig. 6a**). For this, we assessed total *S-Rbm* read count distribution in each run to identify high quality samples. Across the six clinical runs, we assessed the *S-Rbm* sequences further by plotting the number of variant sequences as a function of distinct read percentage cutoffs for each run (**Fig. 6a**). This showed a biphasic distribution with background sequence noise, likely caused by sequencing and PCR errors, increasing exponentially at lower cutoffs. Based on these distributions we assigned run-specific thresholds that minimized background noise, with observed cutoffs ranging between ∼0.16-0.31% (**Supplementary Fig. 12a**). Overall, individual pQS represented less than 5% of the reads for their respective amplicon in both replicates of the barcode analysis for *S-Rbm* and *S-Pbs* (replicate 1: mean = 0.376% and standard deviation = 0.676%; replicate 2: mean = 0.311% and standard deviation = 0.296%; **Supplementary Fig. 13**), in accordance with previous reports that quasispecies are found at low frequencies^22^. We also assessed pQS in non-barcoded *S-Pbs* reads. Here, the background cutoffs ranged between ∼0.127-0.25% (**Supplementary Fig. 12b**) and 1.89% of the tested samples had sequences passing the cutoffs (**Supplementary Fig. 12c**).

**Fig. 6.**
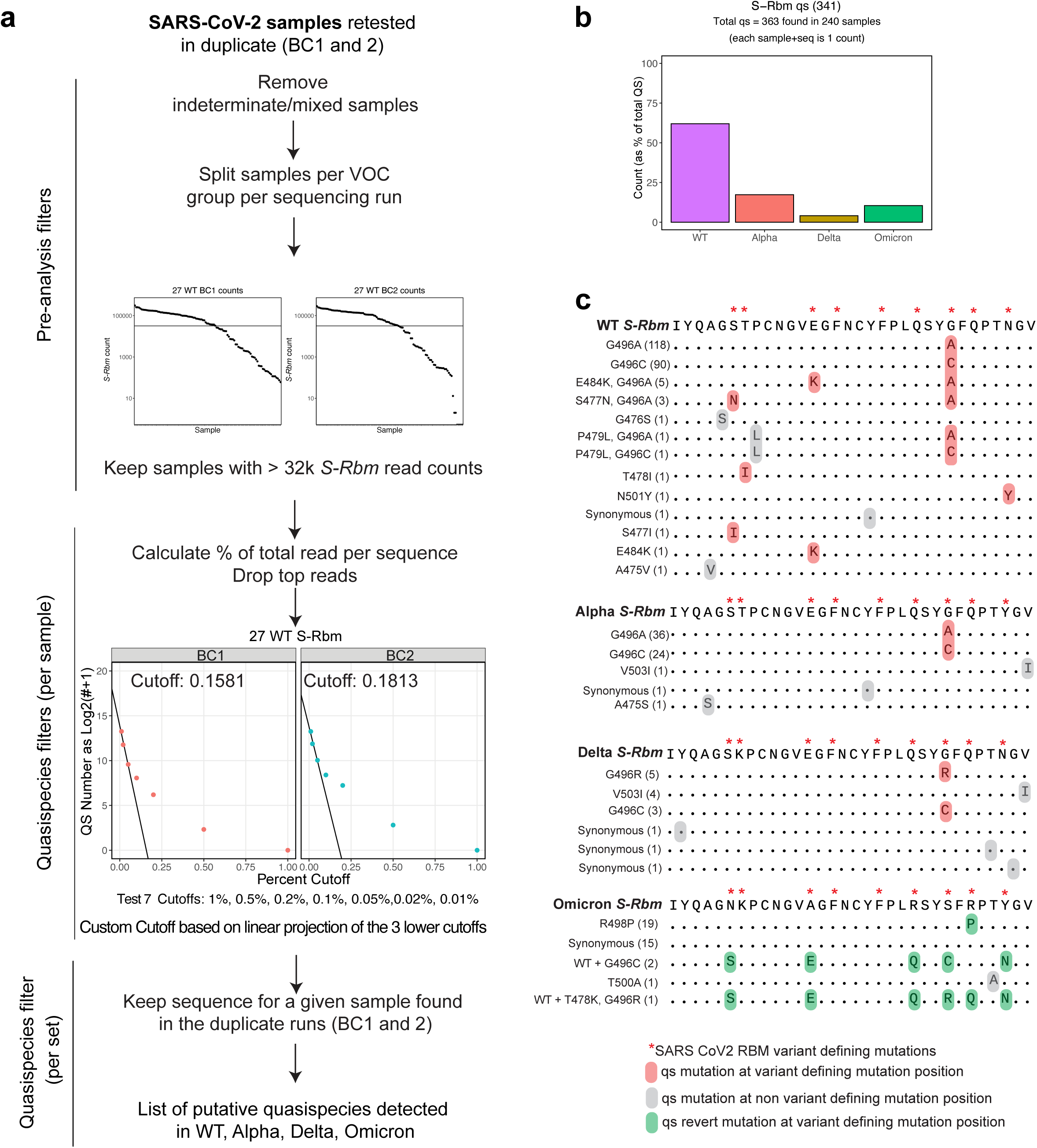
Detection of putative quasispecies by C19-SPAR-Seq. **a.** Schematic of pQS detection. **b.** Percentage of putative quasispecies detected in *S-Rbm* and *S-Pbs* sequences on WT, Alpha, Delta, and Omicron B.1.1.529 samples. **c**. *S-Rbm* and *S-Pbs* putative quasispecies sequences.

Applying these stringent cutoffs to samples across the four tested VOCs yielded a total of 228 samples with *S-Rbm* pQS (20.5% of samples). WT samples showed the highest proportion of *S-Rbm* pQS (65.98%; **Fig. 6b**), while Alpha, Delta, and Omicron accounted for lower (18.48%, 4.39%, and 11.14%, respectively). Of the *S-Rbm* pQS, we identified 29 distinct species, while pQS frequency and number of distinct species was much less for *S-Pbs* (21 samples and 8 species, respectively; **Supplementary Fig. 12d**). For example, the G496A pQS in *S-Rbm* was found in 118 different WT samples (**Fig. 6c**). We identified several samples that possessed multiple pQS in *S-Rbm* (109) or *S-Pbs* (1) and 9 samples with pQS in both *S-Rbm* and *S-Pbs*. Quasispecies analyses suggest that these sequences are not random in nature, but rather cluster around specific mutational profiles. In particular, WT and Alpha were enriched for G496A/C species that were observed in 96.89% and 95.24% of total *S-Rbm* pQS sequences, respectively, but these changes were only in 20% of Delta pQS, where G496R (33.33%) was predominant (**Fig. 6b**). These highlight G496 as an evolutionary target, and indeed G496S became a defining mutation of the Omicron lineage. Similarly, of the other pQS identified in WT, 5 out of 9 variant defining positions in *S-Rbm*, and one variant defining position in *S-Pbs* were targeted that would later be mutated in future VOCs (**Fig. 6c, Supplementary Fig. 12d**, red stars). We also noted that pQS were much more frequent in the *S-Rbm* compared to *S-Pbs* (29 unique sequences *versus* 8), consistent with the key role of the *S-Rbm* in transmission and immune evasion. Interestingly, analysis of pQS in Omicron BA.1.1.529 showed only various combinations of reversion to WT sequences (**Fig. 6c**), suggesting that the Omicron *S-Rbm* is highly optimized for transmission (*ie* ACE2 interaction and immune escape), and that revertants are a favoured evolutionary trajectory. Indeed, R493Q was a WT revertant in Omicron BA 2/3-4/5 and the *S-Rbm* S496G was a revertant in Omicron BA 2/3-4/5 and XBB.1.5 (**Fig. 6c and Supplementary Fig. 14a**). Furthermore, although we sampled Omicron the most, only 6.1% of Omicron samples contributed *S-Rbm* pQS sequences (**Supplementary Table 7**).

### Mutational variants predict future evolutionary trajectories of SARS-CoV-2

To systematically assess whether changes associated with subvariants and pQS in the population might reflect viral sampling of functionally important residues, we developed a mutational compendium for *S-Rbm* and *S-Pbs* by combining subvariant and pQS data (**Fig 5a, Fig. 6c, Fig. 7a, Supplementary Data 3 and Supplementary Fig. 12d**). This showed that *S-Rbm* was highly susceptible to alterations that often predated VOC emergence, particularly around functional residues (**Fig. 7a, left panel**). For example, T478 was identified in both subvariants and pQS of WT virus and subsequently emerged in Delta and Omicron VOCs as a T478K mutation. Similarly, E484 and N501, both of which were altered in multiple strains, were identified in WT populations prior to VOC emergence. In Alpha, we noted that changes in F486 and F490 that extinguished by mid-2021 (see above, **Fig. 5b**), re-appeared as subvariants of the parental Omicron strain BA.1.529, and later became fixed as F486P and F490S in XBB.1.5, BA.2.86, and FLip that emerged through to late 2023^23^. N481 on the other hand was identified in subvariants of later Omicron strains (BA.2, BA.4/5) and emerged in late 2023, as N481K in the BA.2.86 lineage. Interestingly, while the WT virus tended to sample the ACE2 contact residues such as N501 that contributes to ACE2 affinity, subsequent VOCs displayed expansion of variations to include neutralizing Ab contact regions such as F490 and F491 (**Fig. 7a, upper left panel**). Indeed, recent studies show that while alterations in Omicron significantly impact immune recognition, infectivity is variably impacted and typically reduced^23^.

**Fig. 7.**
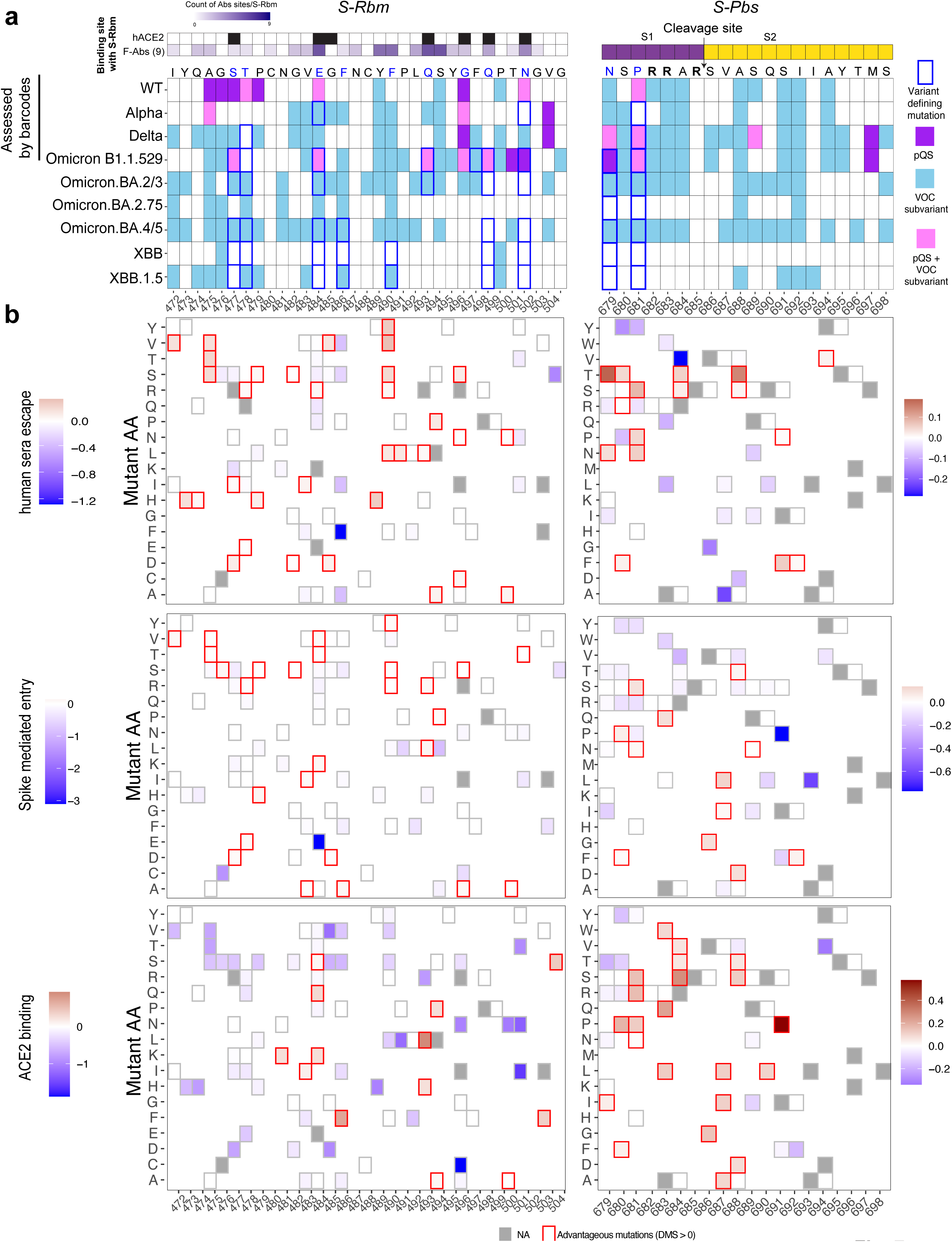
Functional Impact of VOC subvariants and putative quasispecies. **a.** Summary of all mutations and/or putative quasispecies found in *S-Rbm* and *S-Pbs*. *S-Rbm* and *S-Pbs* reference sequences are indicated with VOC-associated changes highlighted in red. At top left, the binding site of *S-Rbm* with hACE2 (black tiles) and 9 neutralizing antibodies are shown (white-purple scale, with darker purple indicating contact with more antibodies)^32, 33, 34^, as well as the *S-Pbs* cleavage site at top right (S1 subunit indicated in purple, S2 subunit indicated in yellow). Changes identified in pQS (purple), subvariants (blue), or both (pink) identified in the indicated VOC are shown. Each variant defining mutations are highlighted in blue for each variant **b.** Functional impact of VOC subvariants and pQS alterations. Alterations identified in systematic screening were compared with functional profiling of Spike deep mutational scanning^25^. Upper panel shows scores obtained for human sera escape, middle panel shows scores from Spike-mediated entry, and lower panel shows scores for ACE2 binding (*S-Rbm* on left, *S-Pbs* on right). For each heatmap, scales for deep mutational scanning scores are shown, with scores above 0 indicating an advantage for viral expansion and highlighted with red outlines. Negative values are blue, indicating no advantage, and are not highlighted. Grey boxes indicate NA values.

We also analyzed the *S-Pbs* that showed mutational sampling of residue N679 and P681 in WT that was subsequently altered in Alpha (P681H), Delta (P681R), Omicron (N679K and P681H), and B.2.86 (N679K and P681R; **Fig. 7a, right panel**). Furthermore, other mutations near the RRxR motif that promote S1/S2 cleavage^24^ were found as VOC subvariants, but did not expand, suggesting they have no major selective advantage for the virus (**Fig. 7a**). Finally, we observed no subvariants or pQS with alterations in R685, which is essential for Furin cleavage. Collectively, these data demonstrate that mapping subvariants and pQS provides predictive and suggest that early viral diversification tends to target viral intrinsic features, while later phases target immune escape.

Our analysis of subvariants and pQS efficiently identified residues mutationally sampled during SARS-CoV-2 viral evolution, but the specific change that emerged in future variants was rarely identified. Indeed, of 158 *S-Rbm* and 87 *S-Pbs* subvariant and pQS changes, only 8 from *S-Rbm* and 3 from *S-Pbs* reflected the specific amino acid alteration that would later emerge (**Fig. 5a, Fig. 6c, Fig. 7a, Supplementary Data 3 and Supplementary Fig. 12d**). Recently, systematic deep mutational scanning (DMS) was performed to measure phenotypes of XBB.1.5 and BA.2 spike proteins, thus providing a map of how all specific mutational changes in Spike impact serum escape, cell entry, and receptor binding ^25^. We took advantage of this study to determine the functional impact of specific *S-Rbm* and *S-Pbs* VOC subvariants and pQS identified by our C19-SPAR-seq pipeline. This showed that most (96%) of observed VOC subvariants and pQS in *S-Rbm* impacted Spike function, while 79% of *S-Pbs* alterations targeted residues with DMS scores (**Fig. 7b**). Many of these changes were measured as advantageous to virus (DMS > 0) for human sera escape (36% of *S-Rbm* and 24.3% of *S-Pbs*; **Fig. 7b, top panels, red tiles**); Spike-mediated entry (28% of *S-Rbm* and 18.6% of *S-Pbs*; **Fig. 7b, middle panels, red tiles**), whereas alterations in S-Rbm targeting ACE2 binding were much less frequent (13%) compared to 31.4% for *S-Pbs* (**Fig. 7b, bottom panels, red tiles**). These studies highlight that the extensive sampling of evolutionary space by SARS-CoV-2 leads to subvariants and pQS changes that are of relevance for specific viral functions.

## Discussion

Population-scale surveillance is crucial to monitor SARS-CoV-2 spreading and its evolution over time. We took advantage of the high-throughput C19-SPAR-seq pipeline to monitor variant spreading and evolution. We screened over 73,510 SARS-CoV-2 positive samples from December 2020 to March 2023 (**Fig. 1a**) and through next-generation sequencing were able to capture both the major top-reads as well as the minor read sequences per patient. This data provides a wealth of sequence information across *S-Rbm* and *S-Pbs*, two of the most critical regions for viral function and amongst the most mutable within the SARS-CoV-2 genome.

Our initial C19-SPAR-Seq pipeline was designed to detect the presence or absence of SARS-CoV-2 within patients in a high-throughput and multiplexed manner. Here, we reasoned that our methodology was amenable to survey the mutational landscape and evolution of SARS-CoV-2 at a population level without much change to the existing methodology. While much of the original pipeline was performed by hand on small numbers of samples, population-level surveillance would require a much more robust and high-throughput pipeline. Utilization of robotics to automate the multiplexing, barcoding, PCR and sample pooling significantly speeds up the processing and reduces human and/or clerical errors. Additionally, we have automated the bioinformatics analysis pipeline such that the hands-on time from sample acquisition to final data output is less than 2 hours of the 24-hour pipeline (**Supplementary Fig. 2**). This truly allows for a one-day turnaround on patient data which is the gold standard during a global pandemic. While this pipeline is robust for the detection of SARS-CoV-2 variants and mutants as we have highlighted here, we provide it as a highly amenable resource for any potential future viral pandemic.

To highlight the flexibility of our pipeline, we had to change our PCR strategy for the detection of the *S-Rbm* around the time of emergence of the Delta VOC, as our initial primers did not capture the T478 residue that defined Delta (**Fig. 2b**). It was essential that we were quickly and readily able to adapt our C19-SPAR-Seq platform with new primers that generated a longer amplicon allowing us to capture this key residue. Importantly, we implemented this change in only four days and tested the assay performance in four independent clinical runs (**Supplementary Fig. 2,3**). The speed at which we were able to implement these changes is critical when faced with highly mutable viral pathogens. While we have focused our analysis here on the *S-Rbm* and *S-Pbs* amplicons, the design and implementation of primers to other regions of the viral genome could easily be added and multiplexed into our assay pipeline seamlessly.

One concern with large amounts of PCR-based NGS is the potential cross-contamination of patient samples and subsequent calling of false positives and/or negatives. We confirmed that the contamination rate of our pipeline was very low (< 3%) by designing the *S-Rbm* V2 primers with an additional barcode (**Supplementary Fig. 5**). The use of these new primers further supports how flexible our assay is to the implementation of new primer pairs. One potential concern about the use of these barcoded primers is that they can become quite costly to use for daily surveillance and monitoring; however, based on the high confidence of correctly mapped reads and well-barcode pairs in our pilot tests (**Supplementary Fig. 2,3**), we opted to use non-barcoded primers for the majority of the daily surveillance with the idea that they could be used if necessary to detect any potential contamination or to assay pipeline performance in sporadic sequencing runs.

Within our jurisdiction we were able to report the dynamic of variant spreading that mirrors the worldwide global SARS-CoV-2 evolution and identified the main VOCs as well as major mutations with these VOCs (**Fig. 2**). Interestingly, we always observed the same repetitive pattern within the population where a VOC would arise as a dominant peak, followed by VOC subvariants that displayed minimal spread, extinguished and were then superseded by the next major incoming VOC (**Fig. 2**). Importantly, by comparing with DMS we found that most subvariants and pQS led to functionally relevant changes, suggesting that population-level spread is mediated by complex interactions that may not be readily predicted from assays of individual viral functions. Integrating how individual molecular functions combine to enable population-level spread could be an important tool for predicting which novel viruses and or their variants are a public health concern.

Through systematic tracking of variant introductions into the GTA, we noted that spread occurred in waves, raising questions about what mechanisms might underlie these dynamics. NPIs could serve as one mechanism for mitigating disease spread within a population, but we failed to note any specific pattern associated with acceleration waves, suggesting NPIs did not make major contributions to regulating the spread of SARS-CoV-2 within our jurisdiction. Similar conclusions with respect to the impact of school closings have been reached^26, 27^, while analysis of viral spread in New York City suggests local geographic constraints might also contribute to this pattern^14^. Furthermore, human social networks display substructure with peak degrees of interaction of approximately 150^28^. This substructure, possibly modified by geographic considerations, vaccination, and prior exposure, may thus collectively create social layers that act to blunt viral spread. Importantly, since current mathematical models, such as the SIR model^29^, assume homogeneity in social networks, they may not be accurate models of viral expansion within the population. In the future it would be of interest to investigate how each of these factors might affect wave dynamics and potential impact of pathogenic viruses in large populations.

Analysis of the major VOC mutations revealed that the initial WT Wuhan strain and subsequent VOCs often gave rise to subvariants with acquired mutations in residues that often became defining features of later VOCs (**Fig. 3,4, and 7**). Furthermore, approximately 25% of our sequencing reads for any given sample did not belong to the predominant sequence and do not constitute contamination (**Supplementary Fig. 5**). For RNA viruses, quasispecies represent the low abundance sequences and heterogeneity found within a patient sample that may lay the foundation for the emergence and evolution of new mutants^18, 19, 20, 21, 22^. We therefore developed a stringent filtering strategy that employed well bar-coding to identify pQS from these high-quality samples and found repeated sequence patterns associated with specific VOCs, consistent with QS theory. This identified 29 unique pQS in *S-Rbm* and 8 in *S-Pbs*, with certain targeted residues that would eventually be mutated in future VOCs. By combining subvariant and pQS monitoring, saily real-time surveillance could thus seed a predictive diagnostic platform that learns the future evolutionary trajectory of highly infectious and/or virulent viruses. This could be an important arm of a One Health strategy that seeks to profile mutational dynamics of high risk viruses with zoonotic potential, such as highly pathogenic avian influenza virus.

In summary, we have developed and optimized our previously reported C19-SPAR-Seq pipeline and deployed it to profile 73,510 SARS-CoV-2-positive patients collected from December 2020 to March 2023 within our jurisdiction. We utilized this platform to identify VOCs, characterize VOC expansion patterns, and prospectively identify mutations characteristic of future evolutionary trajectories. Our development of a systematic, high intensity, relatively low-cost sequencing platform could serve as a blueprint for NGS-based surveillance of other highly mutable pathogens at risk of evolving large-scale transmission characteristics.

## Methods

### Samples collection, Total RNA extraction, and control samples

Positive patient samples were obtained from the Department of Microbiology at Mount Sinai Hospital under MSH REB Study #21-0099-E - Mapping the Emergence and Functional Impact of Novel SARS-CoV-2 Variants. RNA from positive samples was extracted with MGIEasy Nucleic Acid Extraction Kit. We designed and generated the four S gene RNAs to use as an internal control for each C19-SPAR-Seq run or as a matrix for well-BC-S-Rbm-V2 primers. pcDNA3.1 containing the S gene was purchased from Synbio-Technologies. Site-directed mutagenesis was performed to introduce silent mutations into the amplicon region of each gene using the indicated primers and PCR with KOD followed by DpnI (NEB) digestion of the wild-type template DNA (see **Supplementary Data 4**). Successful mutagenesis was confirmed by Sanger sequencing. Single-stranded RNA of each mutant was subsequently produced by *in vitro* translation using the MEGAscript T7 kit (Invitrogen). The four S mutants were used sequentially in the C19-SPARseq as technical control.

### C19-SPAR-Seq primer design and optimization

C19-Spar-Seq used optimized multiplex PCR primers for SARS-CoV-2 (S, *N,* and *RdRP*) and human *ACTB* genes with amplicon size > 100 bases (**Supplementary Table 1**). In this study we used *S-Rbm* V2 primers to generate a larger *Rbm* amplicon (Supplementary Table 1) and well barcoded S-*Rbm*-V2 primers to deconvolute inter- and intrawell contamination as well as determine true minor sequencing mutants (**Supplementary Table 4**)

### Master Mix plates preparation

Reverse Transcript, Multiplex PCR, and Barcode PCR master mix plates (without enzyme and patient samples) were prepared *ex tempo* in 384 well plates in the pre-PCR station. The barcode primer plates were prepared by STARPlus in the pre-PCR station too.

Enzymes (Reverse transcriptase and Polymerase) were dispensed by Echo 555, and RNA, cDNA of patient samples, and the barcode primers were added by the Biomek NxP *ex tempo*.

### Reverse Transcription (RT)

Total RNA was reverse transcribed using SuperScript™ IV Reverse Transcriptase (Invitrogen) in 5X First-Strand Buffer containing DTT, a custom mix of Oligo-dT (Sigma), and Hexamer random primers (Sigma), dNTPs (Genedirex). We followed the manufacturer’s protocol. Each reaction included: 0.25 μL Oligo-dT, 0.25 μL hexamers, 0.5 μL dNTP (2.5 mM each dATP, dGTP, dCTP, and dTTP), 2 μl 5X First-Strand Buffer, 0.5 μl 0.1 M DTT, *quantum satis* (*qs*) 5.5 μL RNase/DNase free water. Then 0.5 μl of SuperScript^TM^ IV RT (200 units/μl) using Echo 555 robotic and 4 μL purified Total RNA was added by Biomek NxP. Samples were incubated at 25°C for 10’, 50°C for 10’, 80°C for 10’, and then stored at 4°C. cDNA was diluted in RNase/DNase-free water at ⅕ by Biomek NxP.

### Multiplexing PCR

The multiplex PCR reaction was carried out using Phusion polymerase (ThermoFisher). The manufacturer’s recommended protocol was followed with the following primer concentrations: *Spoly* at 0.05μM, *SRbm* at 0.05μM, *RdRP* at 0.05μM, and *ActB* at 0.025μM for C19-SPAR-Seq V1 and *Spoly* at 0.038μM, *SRbm-V2* at 0.1μM, *RdRP* at 0.017μM, and *ActB* at 0.025μM for C19-SPAR-Seq V1.2. For each reaction: 2 μL 5X Phusion buffer, 0.2 μL dNTP (2.5 mM each dATP, dGTP, dCTP, and dTTP), primers, *qs* 5.9 μL RNase/DNase free water, then 0.1 μL Phusion Hot start polymerase and 4 μL of diluted cDNA were added by Echo 555 and Biomek NxP respectively. The thermal cycling conditions were as follows: one cycle at 98°C for 2’, and 30 cycles of 98°C for 15’’, 60°C for 15’’, 72°C for 20’’, and a final extension step at 72°C for 5’ and then stored at 4°C. Primer sequences are listed in **Supplementary Table 1**.

### Barcoding PCR

For multiplex barcode sequencing, dual-index barcodes were used^8^. The second PCR reaction on multiplex PCR was performed using the Phusion polymerase (ThermoFisher). For each reaction: 2 μL 5X Phusion buffer, 0.2 μL dNTP (2.5 mM each dATP, dGTP, dCTP, and dTTP), 2 μL Barcoding primers F+R (pre-mix), *qs* 5.9μL RNase/DNase free water, then 0.1 μL Phusion polymerase, 4 μL of multiplex PCR reaction were added by Echo 555 and Biomek NxP respectively. The thermal cycling conditions were as follows: one cycle at 98°C for 30’’, and 15 cycles of 98°C for 10’’, 65°C for 30’’, 72°C for 30’’, and a final extension step at 72°C for 5’ and stored at 4°C.

### Library preparation and Sequencing

Libraries were pooled by Biomex Fx. Each sample was pooled (7uL/sample) and library PCR products were purified twice with SPRIselect beads ratio 1:1 (beads/library) (A66514, Beckman Coulter). All libraries were sequenced with MiSeq or NextSeq 300 or MiniSeq (Illumina) using 100bp single-end sequencing. well-BC libraries were sequenced using Paired-end with Read 1: 144 cycles and Read 2: 8 cycles.

### Whole Genome Sequencing

Sequencing libraries were prepared from extracted patient RNAs using Qiagen QIAseq SARS-CoV-2 Primer Panel (Cat# 333896) according to the manufacturer’s instructions. Library fragment size was then checked using an Agilent Fragment Analyzer and quantified with qPCR using Collibri™ Library Quantification Kit (ThermoFisher, Cat#A38524500) on a BioRad CFX96 Touch Real-Time PCR Detection System. Quality-checked libraries were loaded onto an Illumina NextSeq 500 running with PE 150 cycles. Real-time base call (.bcl) files were converted to FASTQ files using Illumina bcl2fastq2 conversion software v2.17 (on CentOS 6.0 data storage and computation Linux servers). The genome mapping and variant calling were performed by CLC Genomics Workbench V21.0.1 with QIAseq SARS-CoV2 workflow V1.0. The resulting consensus sequences were then uploaded to Pangolin for lineage assignment.

### Cloning and Recombinant Expression of Variants RBM

SARS-CoV2 variant amplicons were obtained by PCR from cDNA generated from reverse transcribed patient samples collected and sequence verified during the recent SARS-CoV2 pandemic. The wild-type (Wuhan variant), Alpha, Beta, Delta, Omicron, and novel (F490L) variants RBM residues 319 - 596 fused with a human signal sequence at the N-terminal were cloned into a pFastBacI vector modified to contain bacterial Biotin ligase (BirA) recognition motif and poly-Histidine tag to the C-terminus. Constructs were expressed as described^30^. Media was dialyzed against a buffer containing 200 mM NaCl, 40 mM Tris HCl pH 7.5, and 10 mM Imidazole overnight. Dialyzed media was incubated in Nickel NTA beads, washed in a Buffer containing 10mM Imidazole, and RBM-His was eluted with 150mM Imidazole.

### RBM Biotinylation

One mg of variants RBM was biotinylated overnight at room temperature in buffer containing 15mL of 1 mg/mL *E. coli* BirA Biotin Ligase, 100 ml of buffer mix B (25 ml 500mM ATP, 25 mL 500mM MgOAc, 62.5Ml D-Biotin 1mM in 0.5M Bicine made up with 12.5 mL ddH_2_O) and purified on 24 mL size exclusion Superdex-200 Column. Biotinylated RBM protein was concentrated, estimated, and stored away at -80°C until used.

### Determination of SARS-CoV-2 RBM-hACE2 Affinity

The affinity between hACE2 and the SARS-CoV-2 S-RBM were determined using streptavidin (SA) biosensors and the Octet RED96 system (ForteBio Inc., Menlo Park, CA, USA). First, the recombinant RBMs were biotinylated, purified on a size-exclusion column (Superdex S75), and immobilized on the SA biosensors at 10 μg/mL in a working buffer containing 25 mM Tris-HCl pH 7.5, 150 mM NaCl, 0.05% Tween-20, and 0.1% BSA. The biosensors were hydrated in the working buffer for 30 minutes at room temperature prior to RBM loading. The hACE2 was diluted in the same working buffer in a two-fold serial dilution series from 970 nM to 15.2 nM (7 different concentrations). The interactions were measured by incubating the loaded sensors in parallel with the seven concentrations of hACE2 and one with only a working buffer as control. The real-time binding response (Δλ in nanometers, nm) at each concentration was corrected by subtracting the background signal measured in the control sample. The kinetic parameters and affinities were estimated based on a one-to-one binding model with a global fit of the data, using the Octet data analysis software (version 7.1, ForteBio Inc., Menlo Park, CA, USA).

### Data Processing

The fastq files were demultiplexed with bcl2fastq and aligned with bowtie (v 0.12.7 with settings --best -v 3 -k 1 -m 1 -S) and counts were quantified with HTSeq-count (v0.13.5 with settings -f sam -t CDS). Custom python scripts (python v3.9) were developed to identify, track, and quantify key mutations (Supplementary Fig2) in the amplicons within samples and to aggregate the data. In early analysis versions, custom R scripts (R v4.1.1 with packages dplyr and openxlsx) were used to apply a quality control filter based on H2O control total viral amplicon counts. In intermediate versions, the minimum read cutoff was calculated as [(median of the control sample *S-Rbm* amplicon counts) + (2*(median absolute deviation of control sample *S-Rbm* amplicon counts))] or a minimum value of 10, whichever is higher. An *S-Rbm* coverage filter to determine confidence in the top read was specified as a 15 percentage point minimum difference between the proportion of WT reads to the top read variants within a sample (for samples where the top read variant was not WT). In final versions of the processing pipeline, the QC functions were added to the python script, the median *S-Rbm-*v2 viral read count cutoff was expanded to include all H2O, HEK, and NoRT controls, and the *S-Rbm* filters were applied to the *S-Rbm-*v2 amplicon instead. Additionally, the final pipeline version is wrapped into a shell script which executes data processing and QC checks in one command.

### Well-Barcoding Methods

To quantify well-to-well contamination we devised a new set of S-*Rbm* PCR barcodes for the rows (R1) and columns (R2) on a 384-well plate. With minor modifications to our existing V2 analysis pipeline, we added R2 processing to assess pairs of reads. Additional code checks for read pairs and then tracks the percentage of each valid row and column PCR barcode pair. We tested 7 plates and found that for non-control samples across all plates, sequences with the correct unique PCR barcode pair assigned to the well made up, on average, 97% of the *S-Rbm* reads per well.

### Rate of Change of Variant Analysis

To calculate VOC case slopes and quantify the rate of change of slopes, the data aggregated for selected VOCs is subsetted per collection day, then quantified against the daily case total across all VOCs to calculate the daily proportion of each VOC. The time in days since the first case of the VOC is calculated with the lubridate package^31^ and linear models are fitted to the data in sliding windows of 5 days with the lm() function to obtain a slope. Missing data is imputed by taking the average of the previous day and the next day’s values. Finally, the slopes are visualized as a heatmap with the x axis representing the first day of the 5-day window to assess the dynamics of the VOC over time.

### Quasispecies Detection

Using the data with *S-Rbm* PCR well-barcodes, we analyzed viral quasispecies in the samples, split into groups by run ID and variant call. Samples failing QC in the normal pipeline, or without a confident variant call, were excluded. Processing is done per run-variant group with a set of custom python and shell scripts (https://github.com/wrana-lab/SPARSEQ_QUASISPECIES). For each sample, the forward and reverse barcodes are checked to verify that they match the sample’s well. Reads surpassing a minimum length are trimmed to remove barcodes and primers and sorted into groups of unique sequences. The groups are quantified as percentages of the total set of sequences per amplicon and per sample, and the most prevalent sequence is dropped. Sequences passing a minimum count threshold are aggregated for further analysis. A set of seven minimum thresholds (0.01%, 0.02%, 0.05%, 0.1%, 0.2%, 0.5%, 1%) are used to assess background levels of sequences per run-variant group. The resulting counts of pQS from the three least stringent cutoffs are used to fit a linear model to calculate the x-intercept for the run-variant group, which is used for a final round of processing as a customized cutoff. Finally, each set of resulting sequences is compared to the corresponding repeated group, and sequences found in the same sample in both copies of the samples are aligned and presented as finalized quasispecies sequences. Any sample where over 5% of the reads are identified as pQS is filtered out. This process was repeated for *S-Pbs* sequences; however, these sequences were not barcoded, so the barcode-matching portion of the pipeline was not applied.

## Code Availability

All of the code required to reproduce these findings and to perform the C19-SPAR-Seq analysis pipeline is available at https://github.com/wrana-lab/SPARseq.

## Data Availability

Data that support the findings of this study have been deposited in the NCBI Gene Expression Omnibus (GEO) under the SuperSeries GSE231416 (https://www.ncbi.nlm.nih.gov/geo/query/acc.cgi?acc=GSE231416) which contains the following subsets: the main dataset including the sample data table is under GSE224951; the well-barcoded dataset is under GSE231415; and the well-barcoded repeat dataset is under GSE246819.

## Acknowledgments

The authors are grateful to all the people who reported to COVID-19 test centers in Toronto and the GTA.

This work is supported by Canadian Institutes of Health Research (Grant #177705) awarded by L.P. and J.L.W. The authors wish to thank the Network Biology Collaborative Centre Robotics Facility (RRID: SCR_025391) at the Lunenfeld-Tanenbaum Research Institute for the automation of the C19-SPAR-Seq platform. The facility is supported by the Canada Foundation for Innovation and the Ontario Government. The facility is supported by the Canada Foundation for Innovation and the Ontario Government. Work in the Pelletier lab was funded by CIHR Foundation (FDN # 167279) and Krembil Foundation. L.P. is a Tier 1 Canada Research Chair in Centrosome Biogenesis and Function. K.N.A. was supported by Medicine by Design Postdoctoral and H.L. Holmes Postdoctoral Fellowships.

## Author contributions

J.L.W., K.N.A., and M.M.A. designed the study. M.M.A., R.P., M.J., P.M.V., N.D., L.Z., A.A-F., F.Y., and A.A. performed C19-SPAR-Seq experiments. L.C., K.N.A., S.P, K-R.Z, A.O., and S.B performed NGS analysis and established the C19-SPAR-Seq interpretation pipeline. L.C., K-R.Z, A.O., and S.B updated and maintained the code. L.C. aggregated and managed the dataset and performed data analysis. M.M.A. and K.N.A. assisted with the rest of the analysis. K.C. performed sequencing. M.M.A. and M.J. set up the pipeline automation. A.A.O, M.J, and K.C. performed biochemistry experiments on the variant affinity assay under the supervision of J.R. and J.L.W.. T.M. provided access to patient samples, collection of diagnostics information, and assembly of the cohorts. P.Y., J.S, J.B., I.L., and B.G.W. coordinated the access to patient samples and generated WGS data. All experiments were carried out under the supervision of L.P and J.L.W. The manuscript was written by M.M.A., L.C., K.N.A., L.P., and J.L.W. with input from T.M. and J.R..

