## Supplementary information for "Systematic surveillance of SARS-CoV-2 reveals dynamics of variant mutagenesis and transmission in a large urban population"

**Supplementary informations:**

**Supplementary Fig. 1 to 14**

**Supplementary Table 1 to 7**

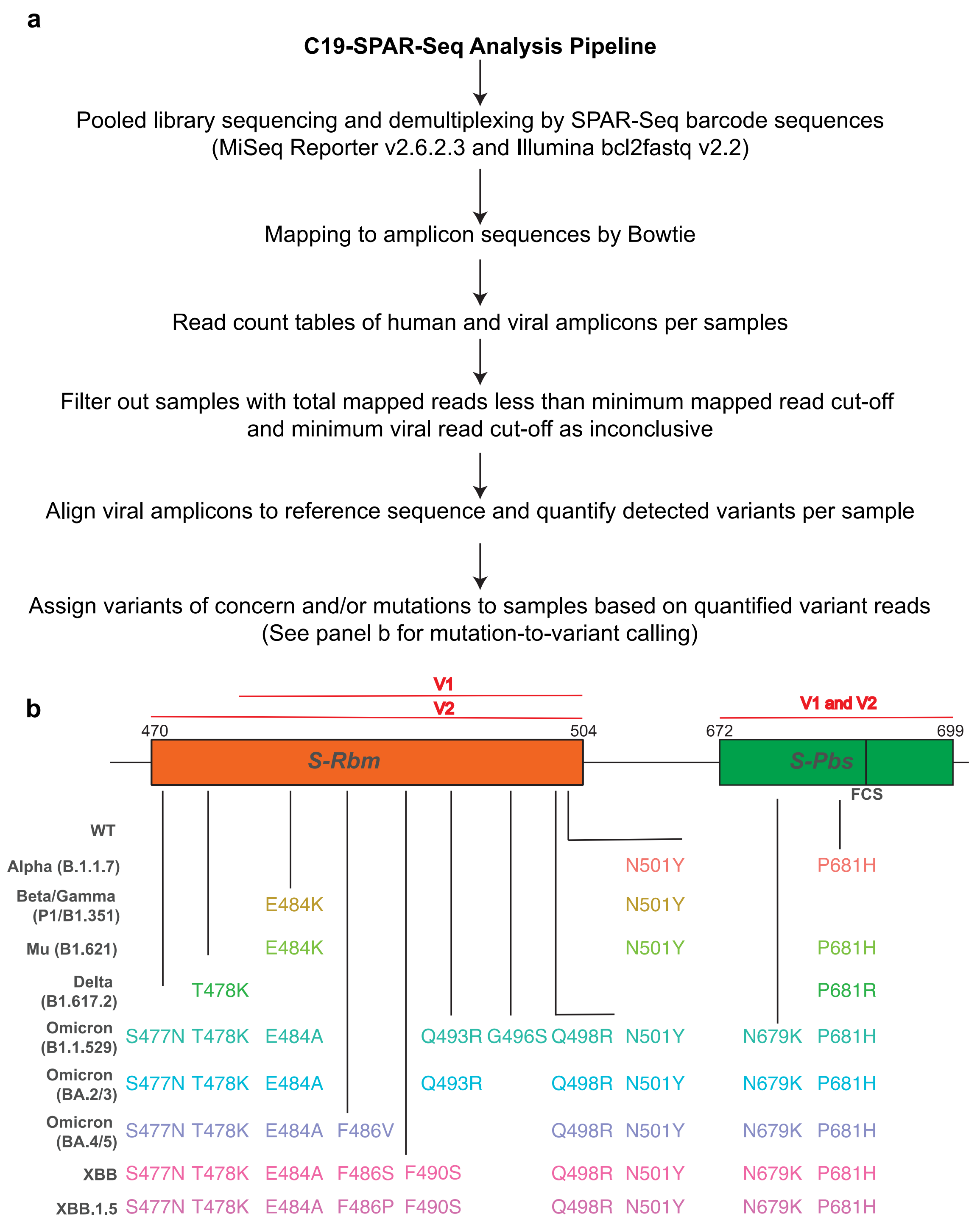


Supplementary Fig. 1

**Supplementary Fig. 1 Summary of analysis pipeline. a.** Each step of the pipeline is detailed. **b.** Schematic of the VOCs and C19-SPAR-Seq V1 and V2 coverage on *S-Rbm* and *S-Pbs*.

*Approx Timeline for one 384 well plate*

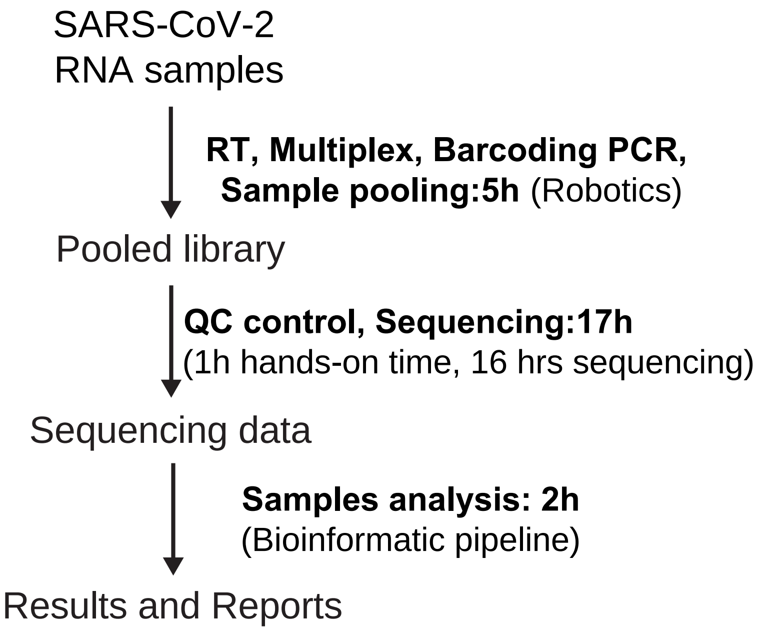


Supplementary Fig. 2

**Supplementary Fig. 2 Timing of the semi-automated C19-SPAR-Seq.**

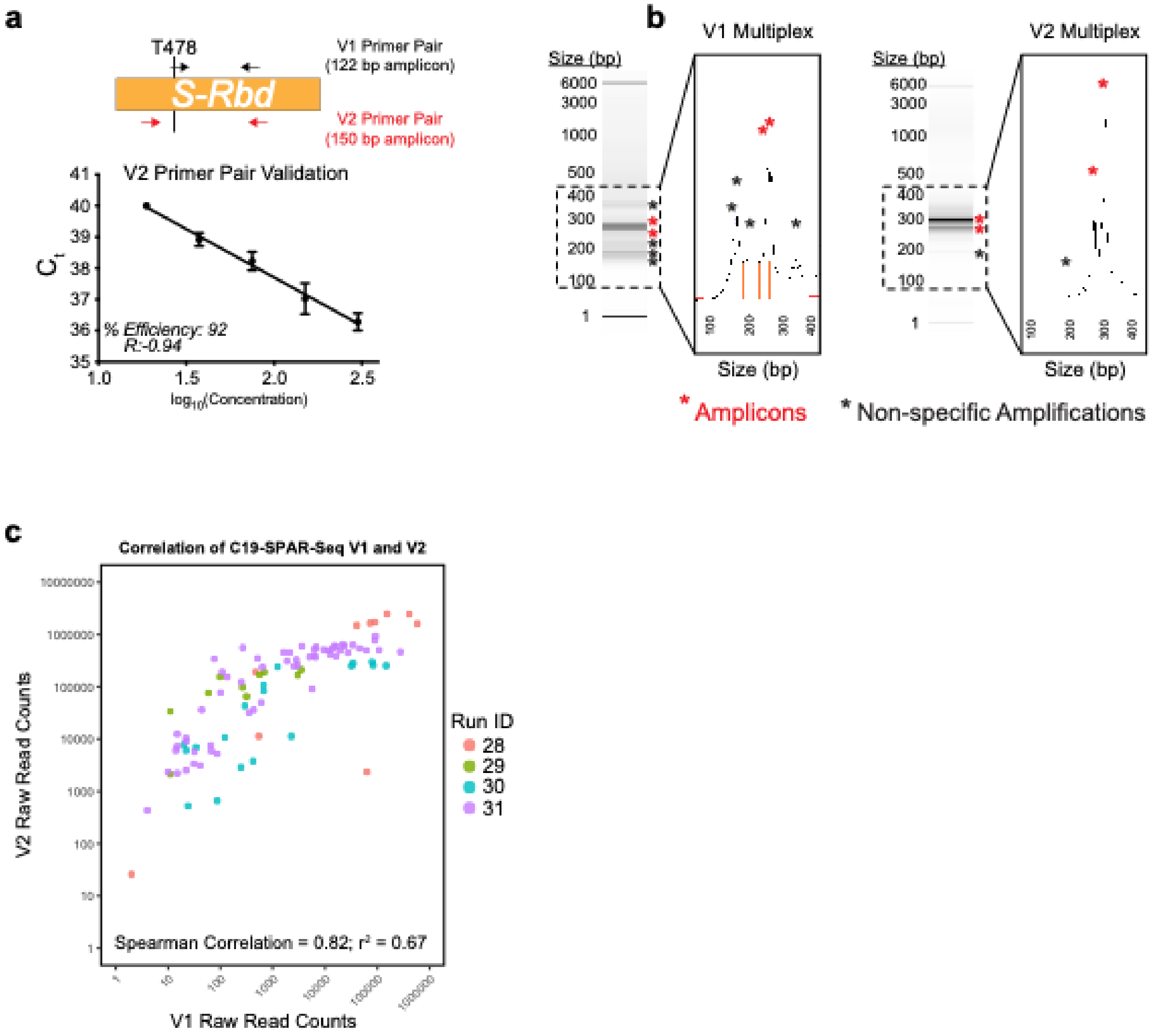


Supplementary Fig. 3

**Supplementary Fig. 3 C19 SPAR-Seq V2 validation. a.** Schematic representation of the V1 and V2 primer pairs of the SARS-CoV-2 *S-Rbm* region (upper panel). The efficiency of *S-Rbm*-V2 set of primers. Standard curve of Ct values (Y-axis) and $\log_{10}$(Concentration) (X-axis) of 5 limited dilutions of HEK293T/Twist RNA. Each condition was tested in duplicate. Means are plotted for each point. After linear regression, the percent efficiency and the Spearman correlation are calculated for each pair of primers (lower panel). **b.** Comparison of fragment analyzer profile of the V1 Multiplex (left panel) and V2 multiplex set of primers (right panel) after 1X/1X SPRI bead purification. Fragment separation (DNA gel) and a blow-up view of the product abundance (electropherogram) are shown. Expected library amplicons (red stars) and non-specific amplicons (black stars) are annotated. **c.** Correlation plot of the total raw read counts of *S-Rbm* using both V1 and V2 primer sets for four clinical runs of the C19-SPAR-Seq pipeline is shown.

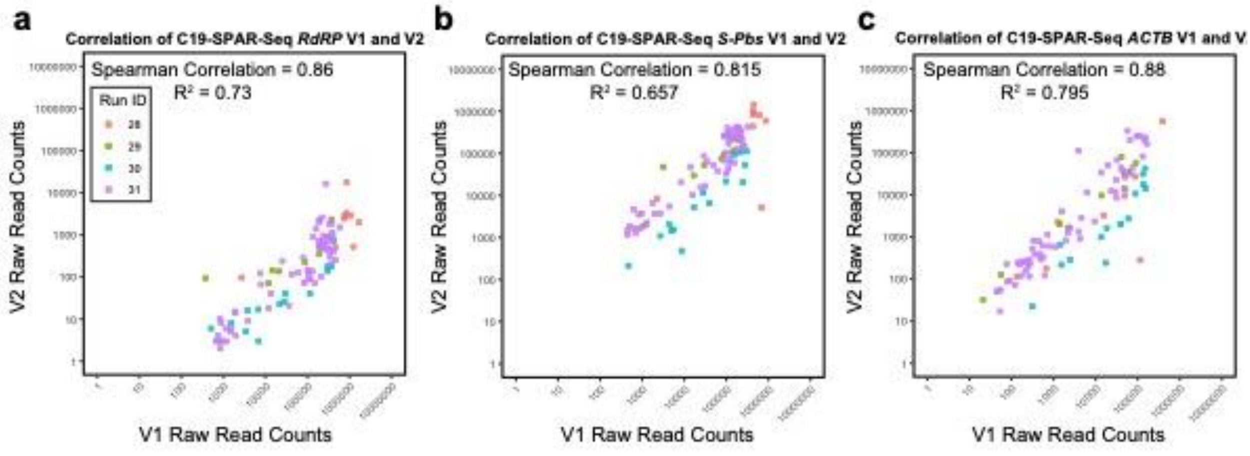


Supplementary Fig. 4

**Supplementary Fig. 4 Correlation plot of the total raw read counts of *RdRP*, *S-Pbs* and *ACTB* using both V1 and V2 primer sets for four clinical runs of C19-SPAR-Seq.**

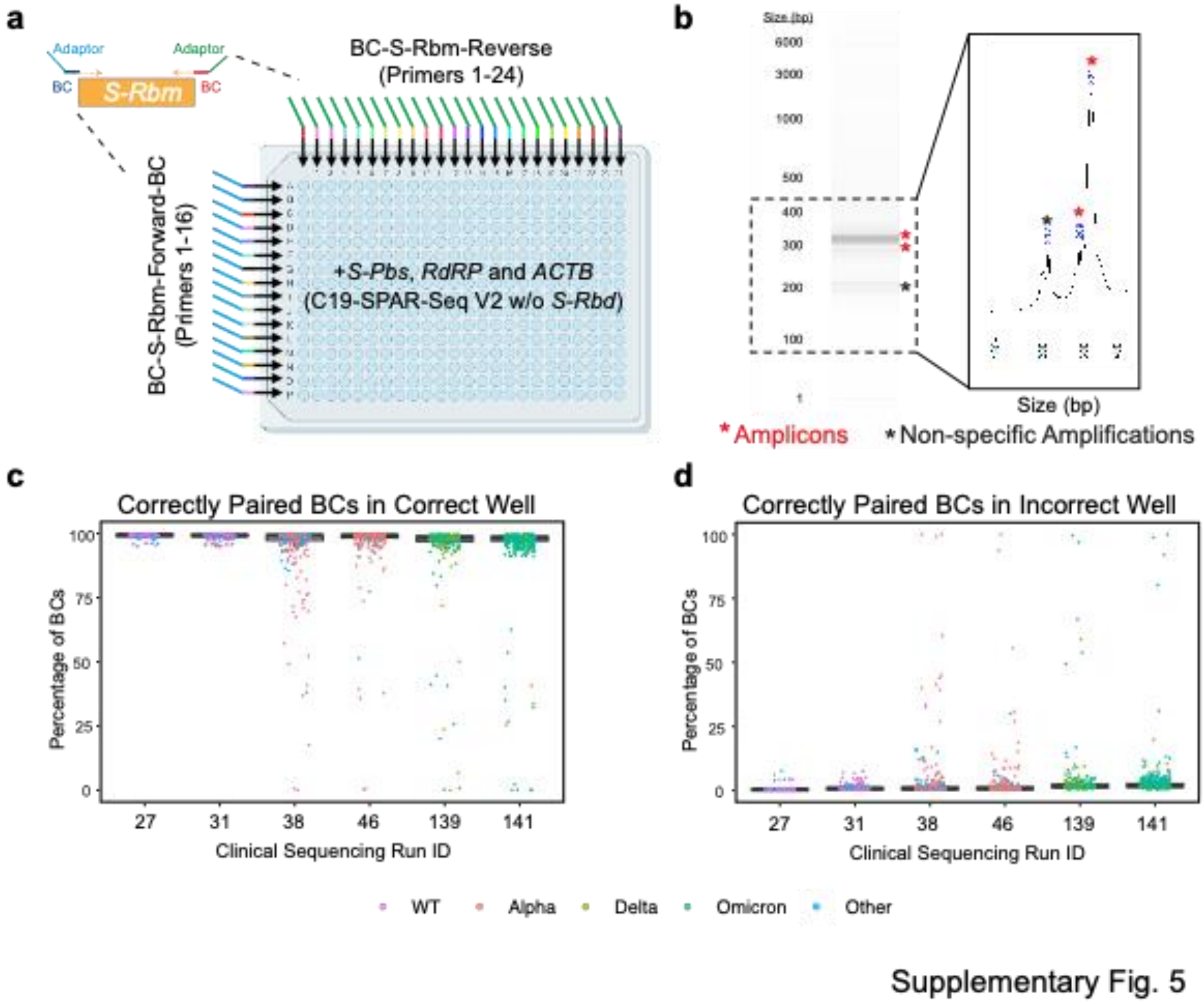


**Supplementary Fig. 5 Quantification of C19-SPAR-Seq technical contamination.** **a.** Schematic of the 384 Well-BC-*S-Rbm* pair of primer designs. **b.** Fragment analyzer profile V2 Well-BC-multiplex set of primers (right panel) after 1X/1X SPRI bead purification. Fragment separation (DNA gel) and a blow-up view of the product abundance (electropherogram) are shown. Expected library amplicons (red stars) and non-specific amplicons (black stars) are annotated. **c.** Boxplot of contamination for the percentage of BCs in the correct well for the six clinical runs. **d**. Boxplot of cross contamination for the percentage of correctly paired BCs in the incorrect well for the six clinical runs.

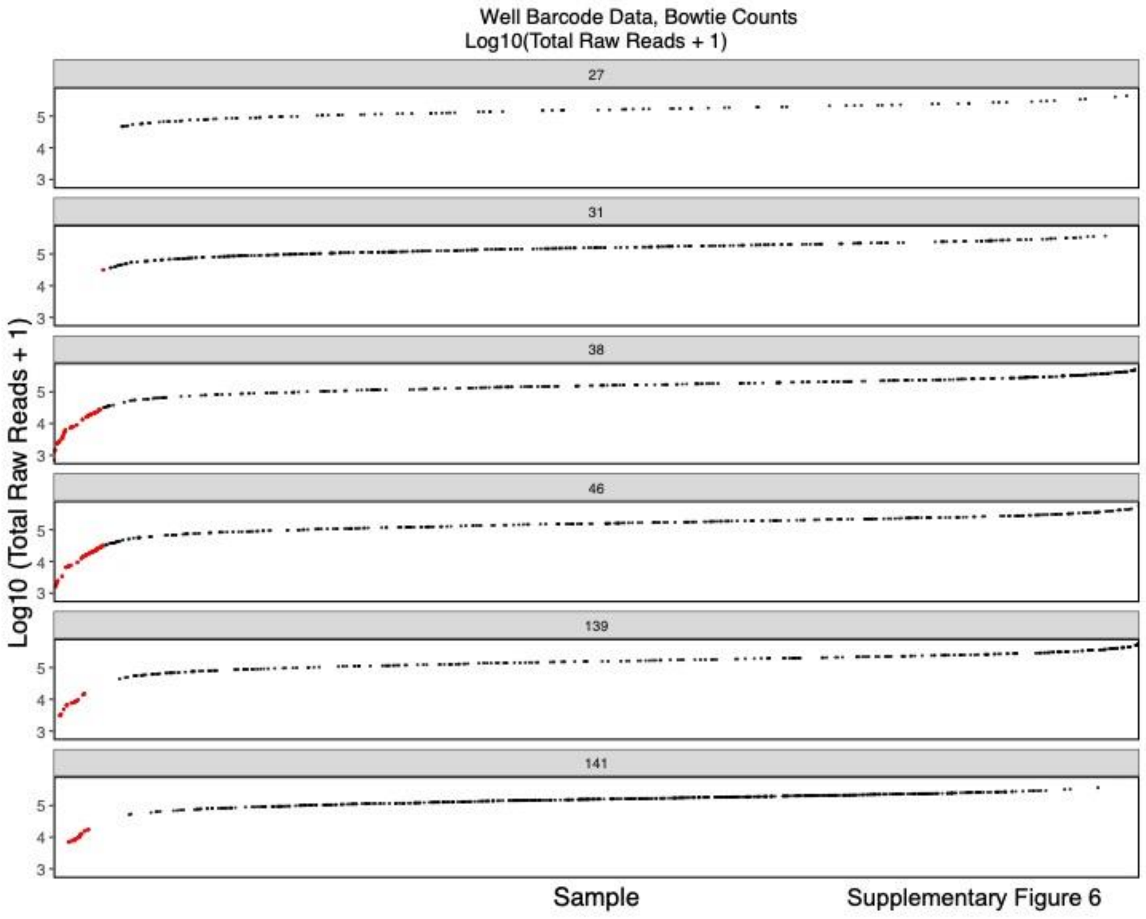


**Supplementary Fig. 6 Total reads for each Barcoded run. Red dots indicate low quality samples.**

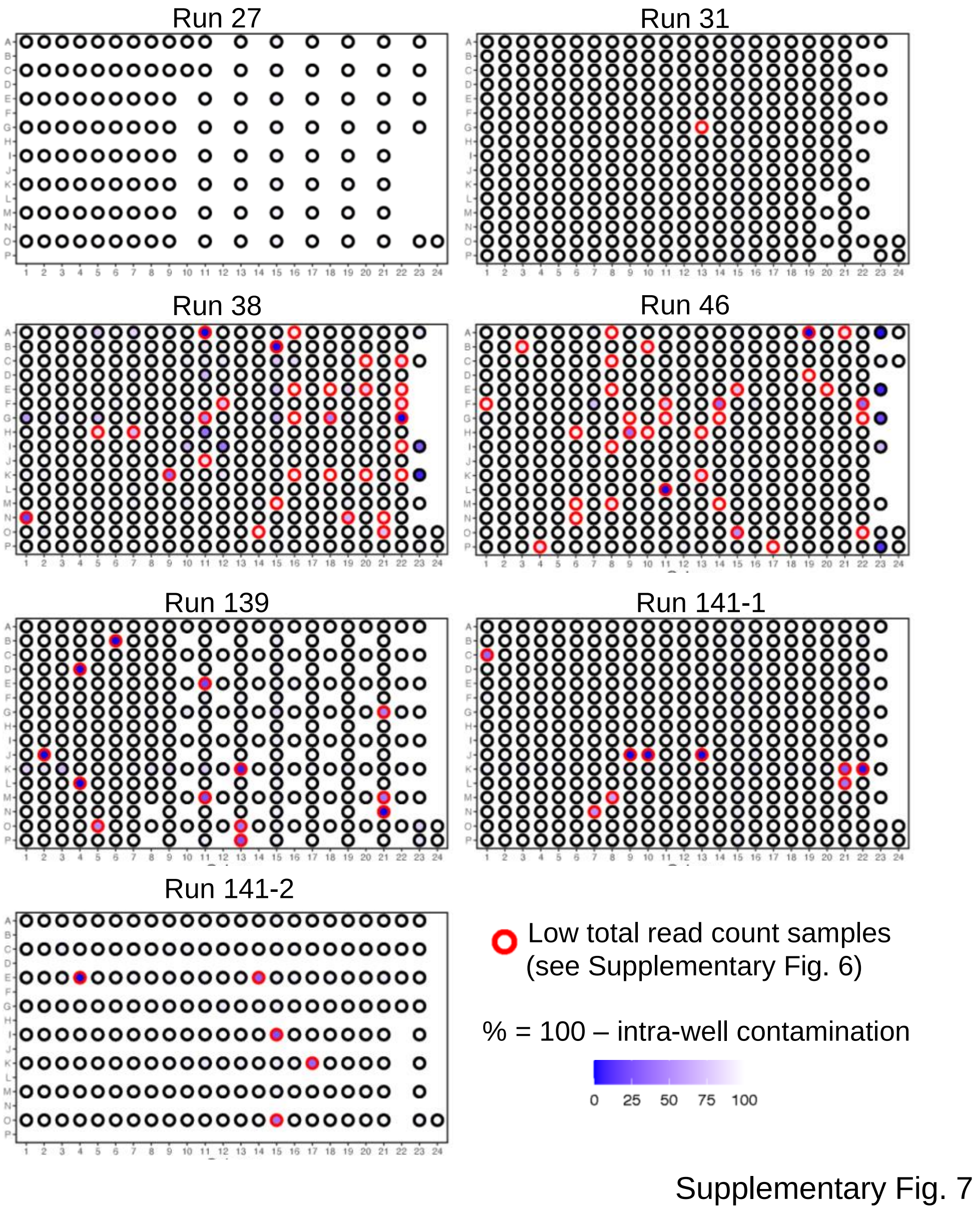


**Supplementary Fig. 7 Schematic of the percentage of correctly paired well-barcoded in the correct well in 384 well plates**. Wells highlighted in red are samples with a low total read count (see Supplementary Fig. 6)

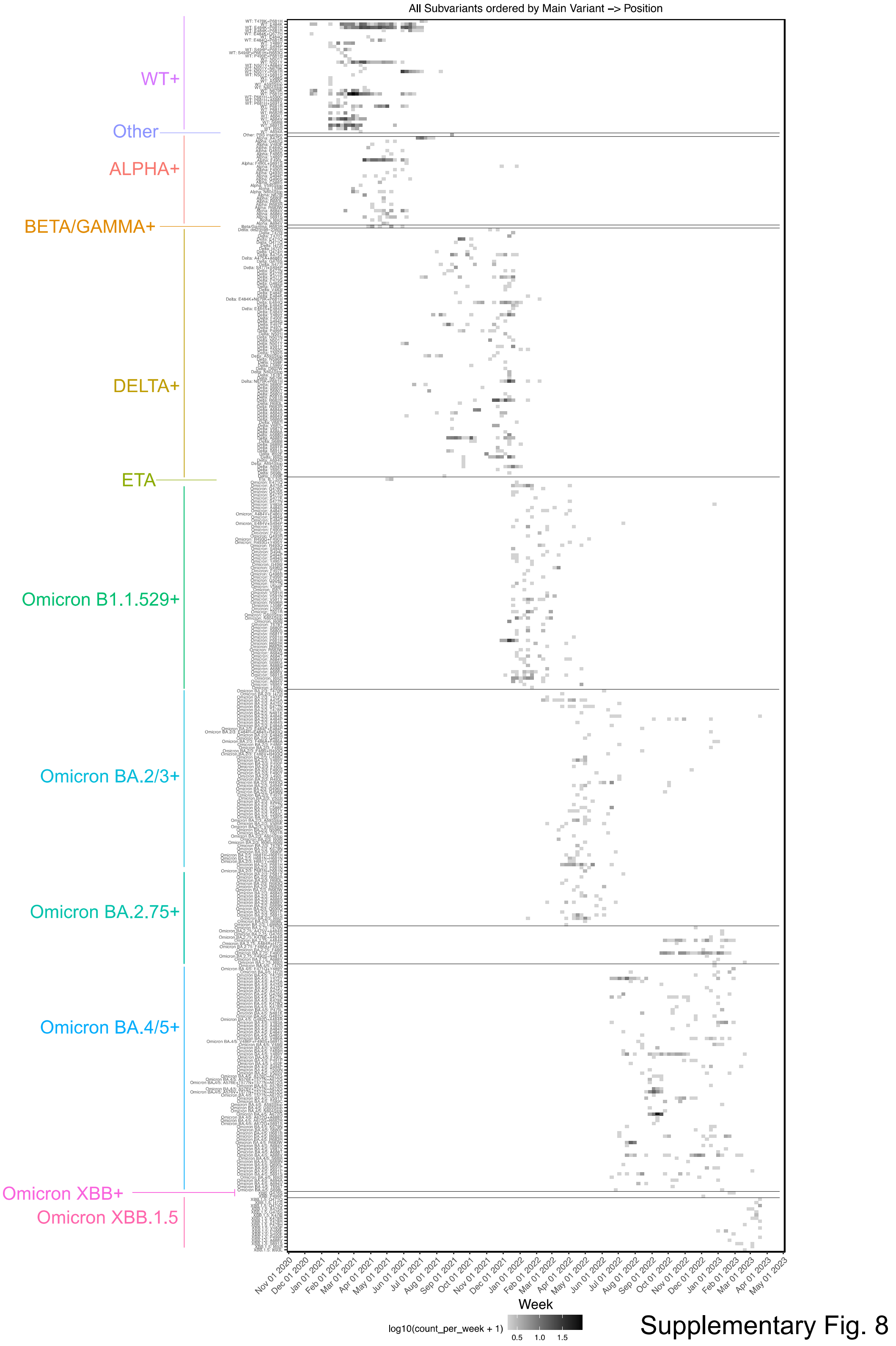


**Supplementary Fig. 8 Heatmap of all the unique VOC subvariants over time.**

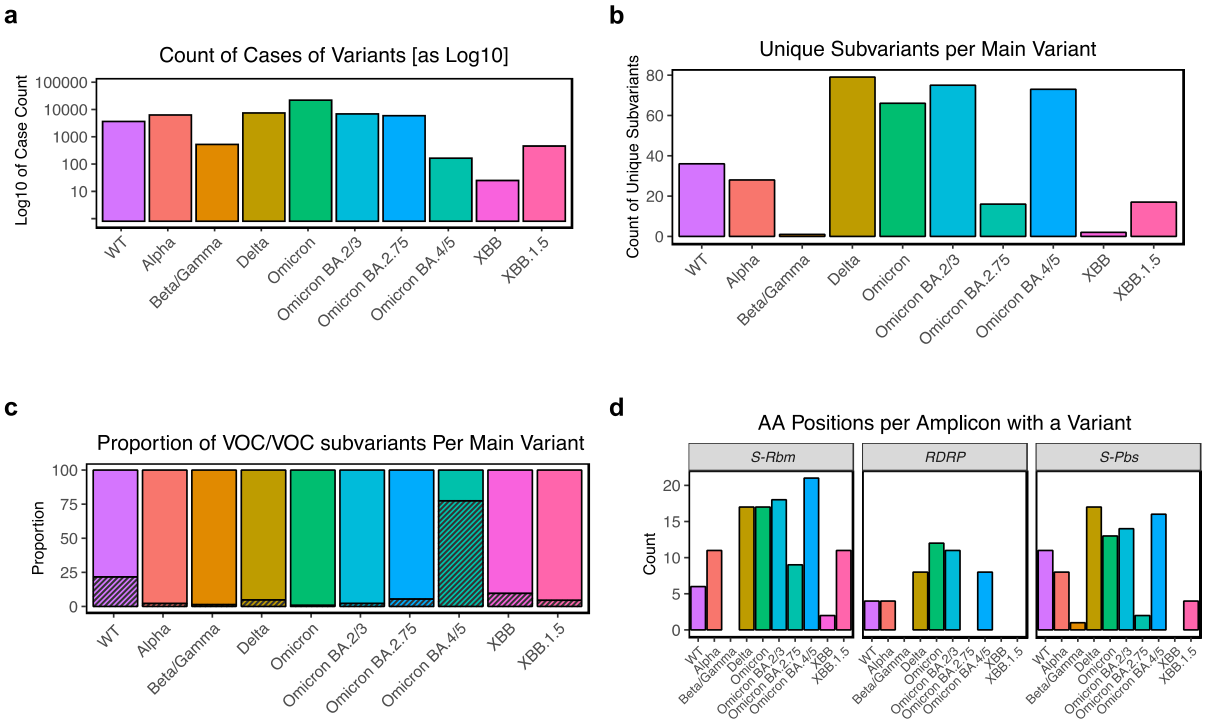


Supplementary Fig. 9

**Supplementary Fig. 9 VOCs and VOC subvariant distribution. a.** Total cases of each VOC. **b.** Total number of different unique mutational profiles per VOC. **c.** Proportion of cases of VOCs with subvariants (shaded areas). **d.** For each amplicon, the number of amino acid positions in which we detected a variant in the different VOCs.

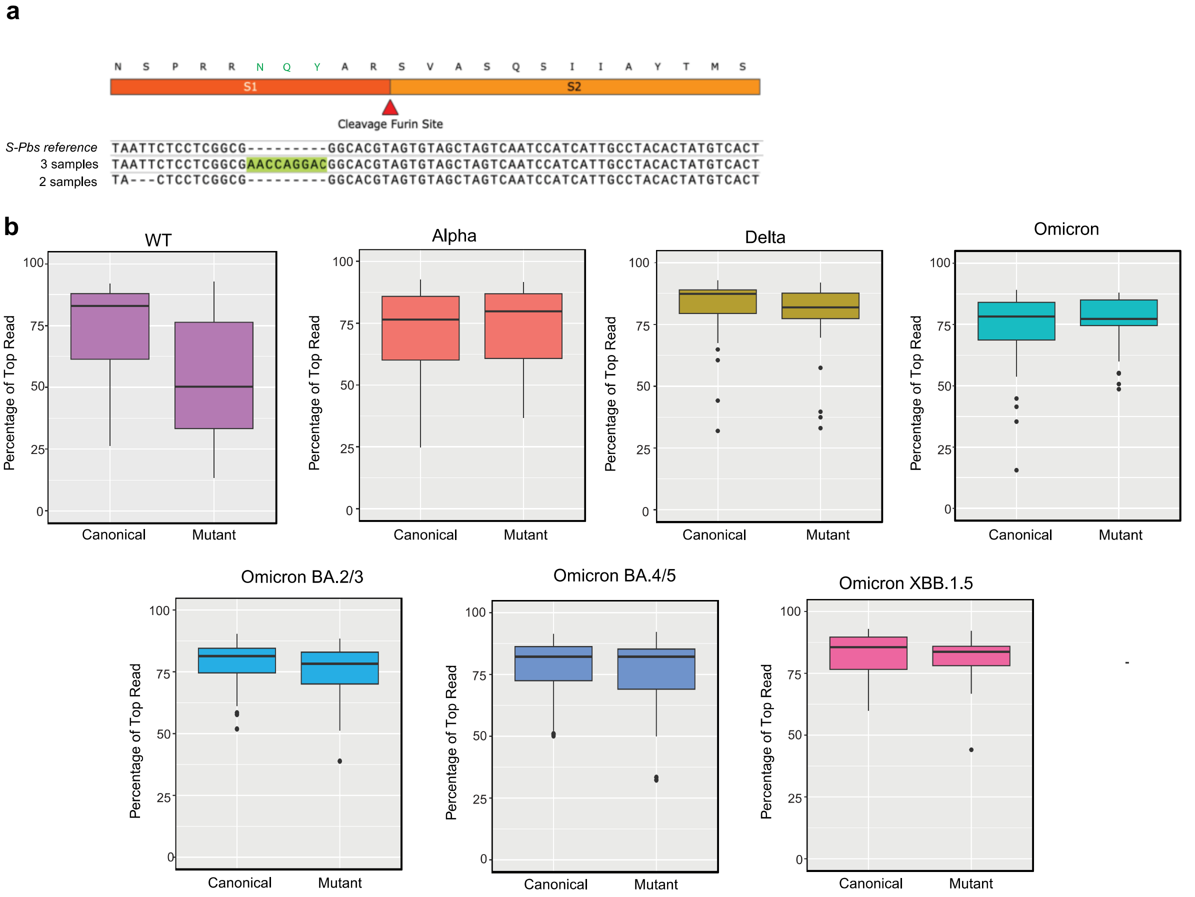


Supplementary Fig. 10

**Supplementary Fig. 10 VOC subvariants. a.** VOC subvariant of *S-Pbs* insertion and deletion. **b.** The percentage of the top reads out of the total read counts per amplicon, per sample, are plotted for each of the VOCs and VOC subvariants.

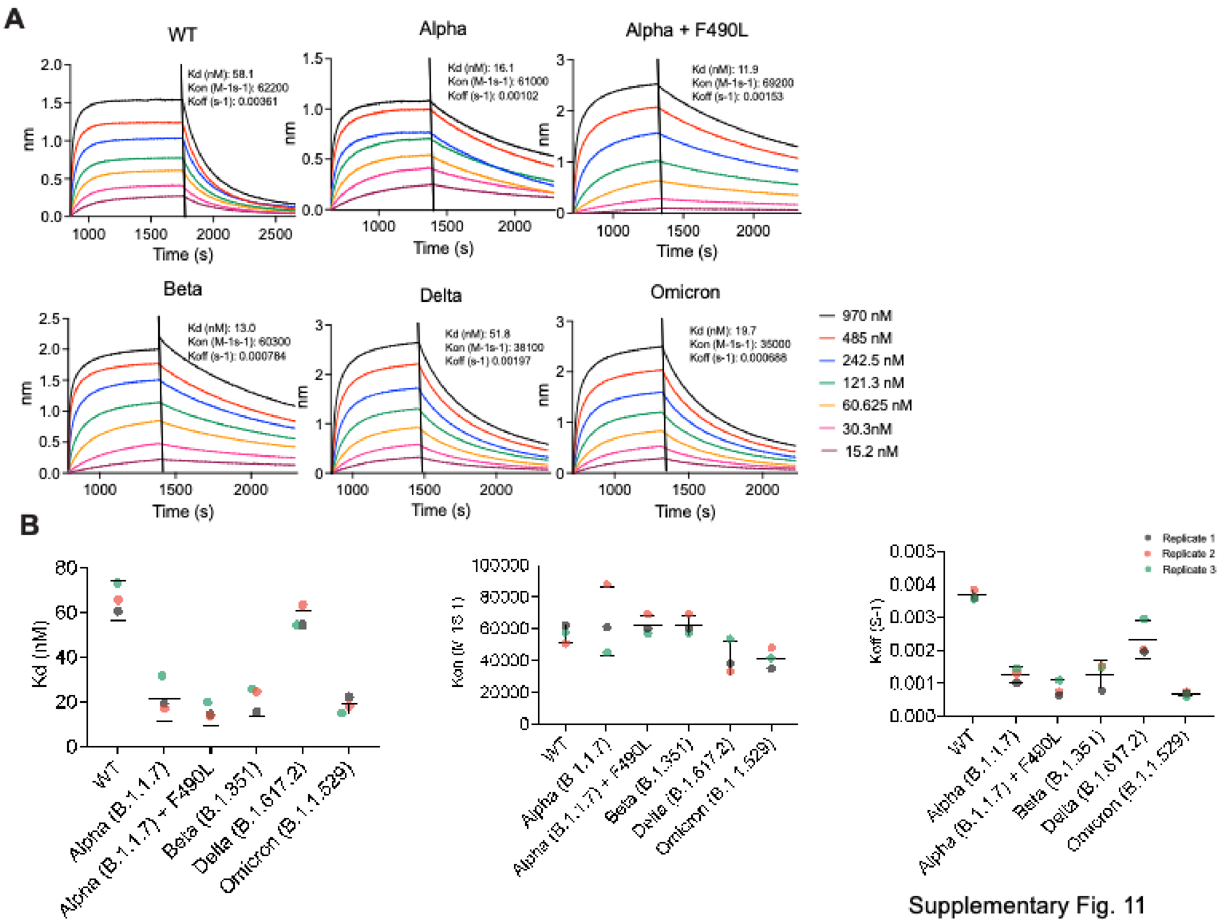


Supplementary Fig. 11

**Supplementary Fig. 11 Affinity Assay of WT, Alpha, Alpha + F490L, Beta, Delta and Omicron**

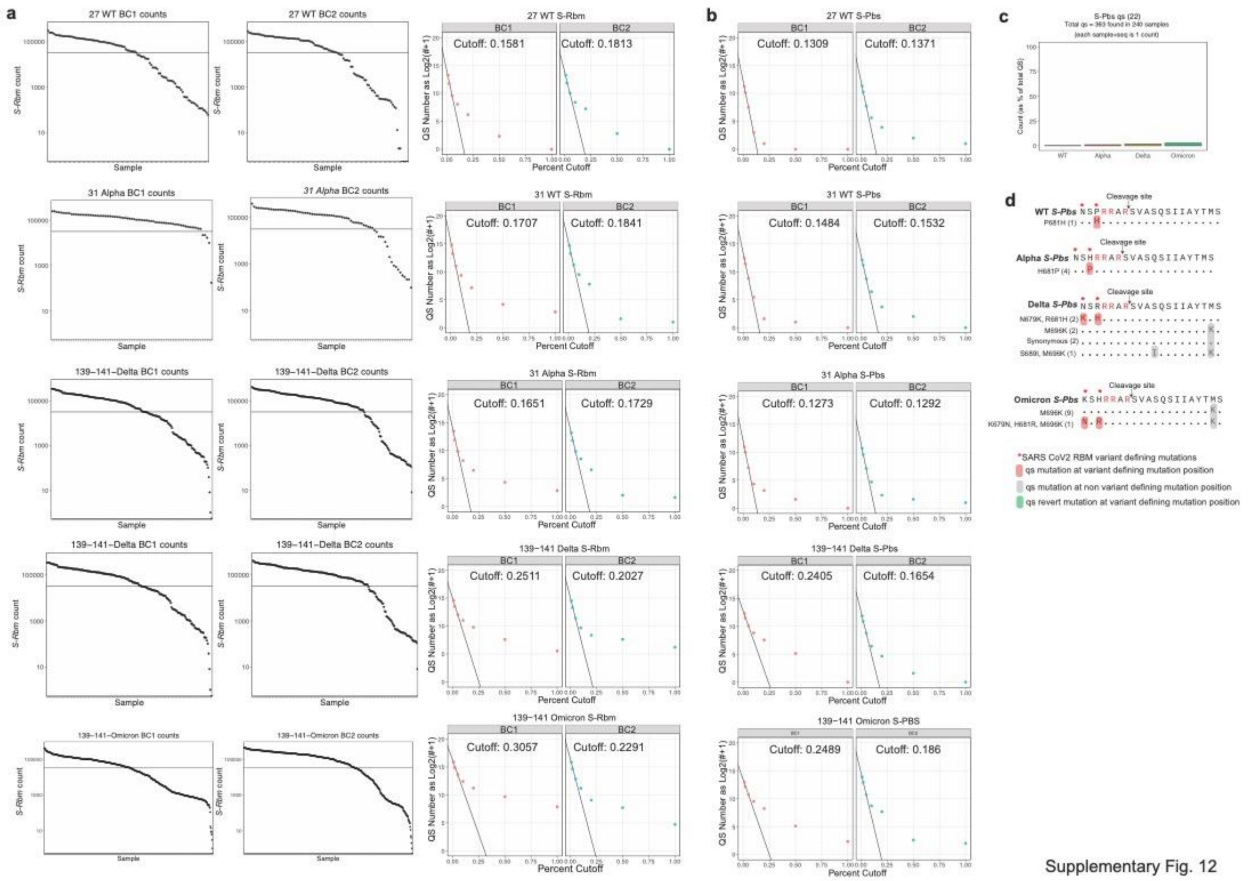

**Supplementary Fig. 12 Filtering and threshold development for putative quasispecies analysis, and pQS results for *S-Pbs* sequences. a.** Counts per sample of *S-Rbm* reads with the cutoff line shown at 32,000 counts (left panel) and analysis to determine the finalized threshold for *S-Rbm* reads (right panel). **b.** Analysis to determine the finalized threshold for *S-Pbs* reads. **c.** Count of *S-Pbs* pQS per VOC as percent of total detected pQS. **d.** Alignment of *S-Pbs* pQS sequences.

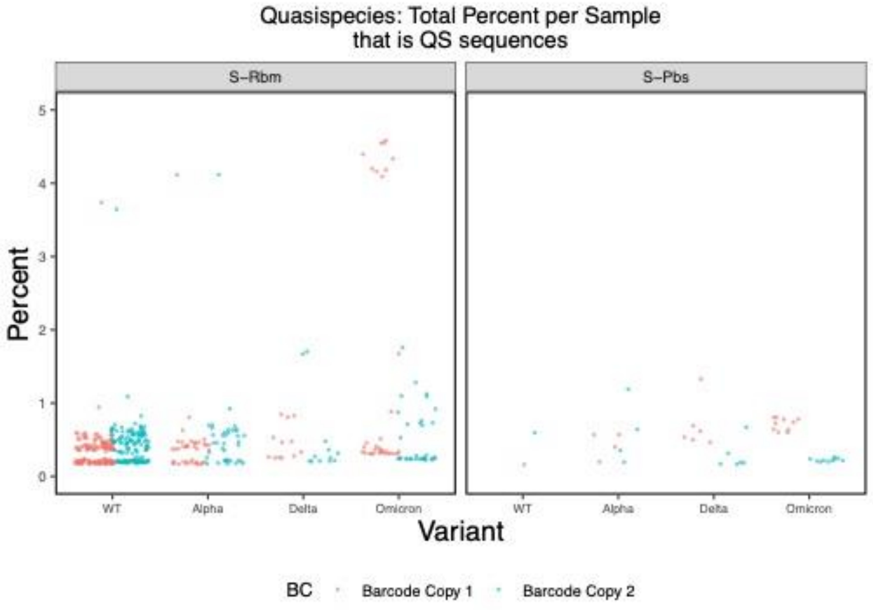


Supplementary Fig. 13

**Supplementary Fig. 13 Total percent of reads per sample that is made up of pQS sequences.**

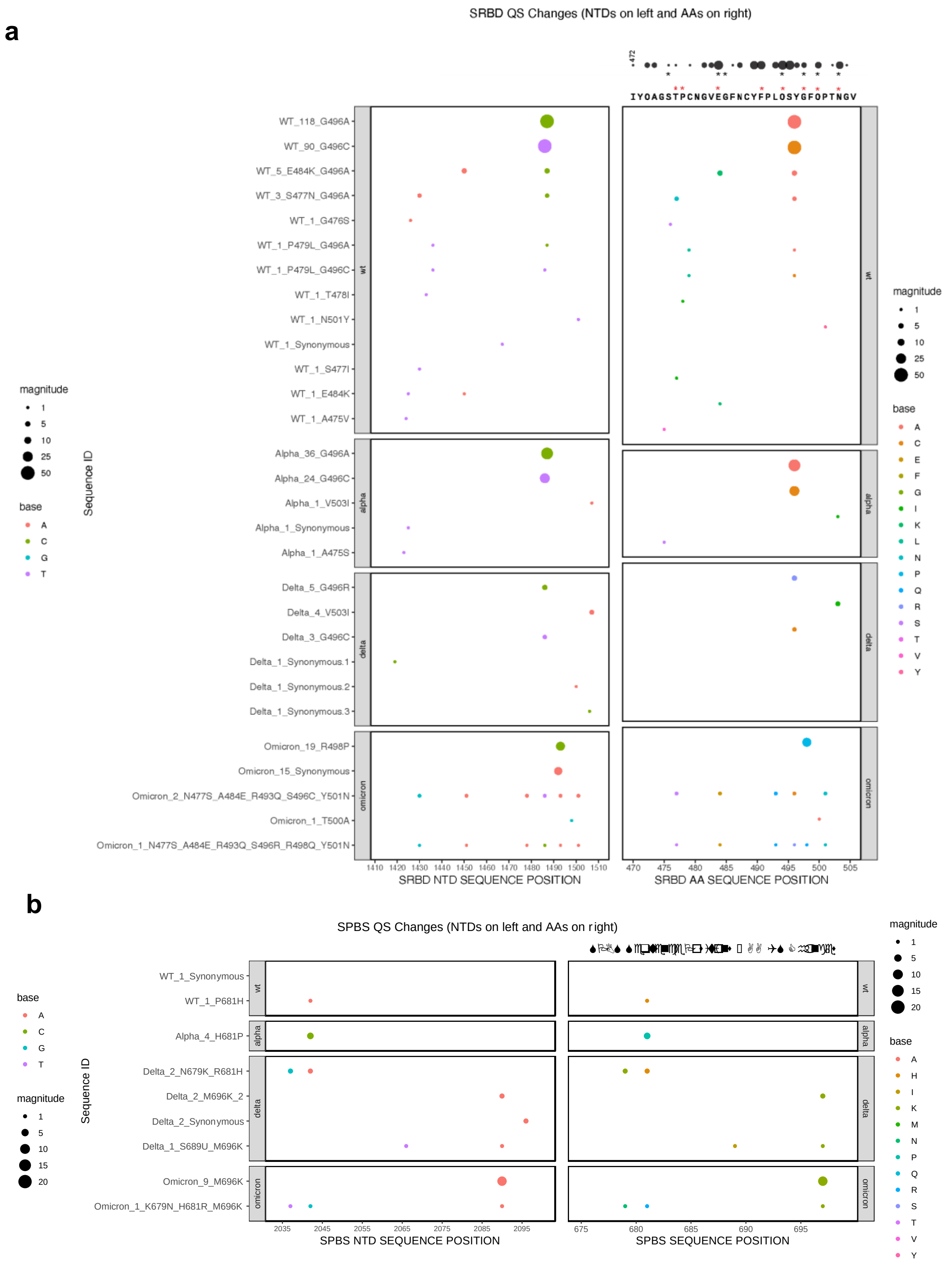


Supplementary Fig. 14

**Supplementary Fig. 14 Summary of *S-Rbm* and *S-Pbs* pQS. a.** *S-Rbm* pQS: nucleotide mutation (left panel) and amino acid mutation (right panel). **b.** *S-Pbs* pQS: nucleotide mutation (left panel) and amino acid mutation (right panel).

| sequence | Name | Version |
|---|---|---|
| acactctttccctacacgacgctcttccgatctATCAGGCCGGTAGCACACCT | 013-S-RBD-For | V1 |
| gtgactggagttcagacgtgtgctcttccgatctACTCTGTATGGTTGGTAACCAACAC | 013-S-RBD-Rev | V1 |
| acactctttccctacacgacgctcttccgatctTATGCGCTAGTTATCAGACTCAGAC | 014-S-PBS-For | V1/V2 |
| gtgactggagttcagacgtgtgctcttccgatctGTAAGCAACTGAATTTTCTGCACCA | 014-S-PBS-Rev | V1/V2 |
| acactctttccctacacgacgctcttccgatctGATGCCACAACTGCTTATGC | 023-RdRP-For | V1/V2 |
| gtgactggagttcagacgtgtgctcttccgatctTTGCGGACATACTTATCGGC | 023-RdRP-Rev | V1/V2 |
| acactctttccctacacgacgctcttccgatctTCACCATTGGCAATGAGCGGTTC | 019-ACTB/G-For | V1/V2 |
| gtgactggagttcagacgtgtgctcttccgatctCCACGTCACACTTCATGATGGAG | 019-ACTB/G-Rev | V1/V2 |
| acactctttccctacacgacgctcttccgatctACCTTTTGAGAGAGATATTTCAACTGA | S-RBD-V2-For | V2 |
| gtgactggagttcagacgtgtgctcttccgatctACTACTACTCTGTATGGTTGGT | S-RBD-V2-Rev | V2 |

**Supplementary Table 1. List of SARS-CoV-2 and human primers**

| Description | Number Failed in Pilot Cohort (from 1,218 sample | Number Failed in Total Cohort (from 73,510 samples) |
|---|---|---|
| QC Fail | 20 | 7,345 |
| QC Pass | 1,198 | 66,165 |

**Supplementary Table 2. Summary of samples which failed quality control cutoffs.**

| Sequence name | Lineage | Conflict | Ambiguity score | Scorpio call | Scorpio support | Scorpio conflict | Note | pangolin version | pangolin-data version | Variant type SPAR-Seq detected |
|---|---|---|---|---|---|---|---|---|---|---|
| W42207758 | B.1.2 | 0 | | | | | Assigned from designation hash. | 4.1.3 | v1.15.1 | C179 (Asn -> Lys) |
| W41900420 | B.1.2 | 0 | | | | | Usher placements: B.1.2(1/1) | 4.1.3 | v1.15.1 | C484 (Glu -> Lys) |
| W41809599 | B.1.2 | 0 | | | | | Assigned from designation hash. | 4.1.3 | v1.15.1 | C179 (Asn -> Lys) |
| W41808799 | B.1.36.18 | 0 | | | | | Assigned from designation hash. | 4.1.3 | v1.15.1 | High |
| W41805394 | AL.1 | 0 | | | | | Assigned from designation hash. | 4.1.3 | v1.15.1 | High |
| W41809402 | B.1.469 | 0 | | | | | Usher placements: B.1.469(1/1) | 4.1.3 | v1.15.1 | C681(Pro -> His) |
| W41809395 | B.1.469 | 0 | | | | | Assigned from designation hash. | 4.1.3 | v1.15.1 | C681(Pro -> His) |
| W41809377 | B.1.243 | 0 | | | | | Assigned from designation hash. | 4.1.3 | v1.15.1 | C681(Pro -> His) |
| W41907070 | P.2 | 0 | | Zeta (P.2-like | 0.86 | 0.14 | Assigned from designation hash. | 4.1.3 | v1.15.1 | C484 (Glu -> Lys) |
| W41907062 | P.2 | 0 | | Zeta (P.2-like | 0.86 | 0.14 | Assigned from designation hash. | 4.1.3 | v1.15.1 | C484 (Glu -> Lys) |
| W41906025 | B.1.2 | 0 | | | | | Usher placements: B.1.2(1/1) | 4.1.3 | v1.15.1 | C179 (Asn -> Lys) |
| W42002970 | B.1.36.18 | 0 | | | | | Usher placements: B.1.36.18(1/1) | 4.1.3 | v1.15.1 | *Medium* |
| W42005505 | B.1.2 | 0 | | | | | Assigned from designation hash. | 4.1.3 | v1.15.1 | C179 (Asn -> Lys) |
| W42004805 | B.1.36.38 | 0 | | | | | Assigned from designation hash. | 4.1.3 | v1.15.1 | *Medium* |
| W42005597 | AM.4 | 0 | | | | | Assigned from designation hash. | 4.1.3 | v1.15.1 | C484 (Glu -> Lys) |
| W42204655 | B.1.2 | 0 | | | | | Assigned from designation hash. | 4.1.3 | v1.15.1 | C179 (Asn -> Lys) |
| W42104455 | B.1.243 | 0 | | | | | Assigned from designation hash. | 4.1.3 | v1.15.1 | C681(Pro -> His) |
| W42205151 | P.2 | 0 | | Zeta (P.2-like | 0.86 | 0.14 | Assigned from designation hash. | 4.1.3 | v1.15.1 | C484 (Glu -> Lys) |
| W42103083 | B.1.36.18 | 0 | | | | | Usher placements: B.1.36.18(1/1) | 4.1.3 | v1.15.1 | Low |
| W42203027 | B.1.2 | 0 | | | | | Assigned from designation hash. | 4.1.3 | v1.15.1 | C179 (Asn -> Lys) |
| W42207759 | B.1.2 | 0 | | | | | Assigned from designation hash. | 4.1.3 | v1.15.1 | C179 (Asn -> Lys) |

**Supplementary Table 3. Pangolin output of whole genome sequencing results.**

| Name | Sequence |
|---|---|
| S_Rbd_V2_For_well-BC_1 | acactctttccctacacgacgctcttccgatct**ACGTA**ACCTTTTGAGAGAGATATTTCAACTGA |
| S_Rbd_V2_For_well-BC_2 | acactctttccctacacgacgctcttccgatct**CGTAC**ACCTTTTGAGAGAGATATTTCAACTGA |
| S_Rbd_V2_For_well-BC_3 | acactctttccctacacgacgctcttccgatct**GTACC**ACCTTTTGAGAGAGATATTTCAACTGA |
| S_Rbd_V2_For_well-BC_4 | acactctttccctacacgacgctcttccgatct**TACGG**ACCTTTTGAGAGAGATATTTCAACTGA |
| S_Rbd_V2_For_well-BC_5 | acactctttccctacacgacgctcttccgatct**AGGAC**ACCTTTTGAGAGAGATATTTCAACTGA |
| S_Rbd_V2_For_well-BC_6 | acactctttccctacacgacgctcttccgatct**CTTCG**ACCTTTTGAGAGAGATATTTCAACTGA |
| S_Rbd_V2_For_well-BC_7 | acactctttccctacacgacgctcttccgatct**GAAGT**ACCTTTTGAGAGAGATATTTCAACTGA |
| S_Rbd_V2_For_well-BC_8 | acactctttccctacacgacgctcttccgatct**TCCTA**ACCTTTTGAGAGAGATATTTCAACTGA |
| S_Rbd_V2_For_well-BC_9 | acactctttccctacacgacgctcttccgatct**AGTGC**ACCTTTTGAGAGAGATATTTCAACTGA |
| S_Rbd_V2_For_well-BC_10 | acactctttccctacacgacgctcttccgatct**CTATG**ACCTTTTGAGAGAGATATTTCAACTGA |
| S_Rbd_V2_For_well-BC_11 | acactctttccctacacgacgctcttccgatct**GATAC**ACCTTTTGAGAGAGATATTTCAACTGA |
| S_Rbd_V2_For_well-BC_12 | acactctttccctacacgacgctcttccgatct**TCACG**ACCTTTTGAGAGAGATATTTCAACTGA |
| S_Rbd_V2_For_well-BC_13 | acactctttccctacacgacgctcttccgatct**AGCGT**ACCTTTTGAGAGAGATATTTCAACTGA |
| S_Rbd_V2_For_well-BC_14 | acactctttccctacacgacgctcttccgatct**CTGAC**ACCTTTTGAGAGAGATATTTCAACTGA |
| S_Rbd_V2_For_well-BC_15 | acactctttccctacacgacgctcttccgatct**GATCG**ACCTTTTGAGAGAGATATTTCAACTGA |
| S_Rbd_V2_For_well-BC_16 | acactctttccctacacgacgctcttccgatct**TCGGA**ACCTTTTGAGAGAGATATTTCAACTGA |
| S_Rbd_V2_Rev_well-BC_1 | gtgactggagttcagacgtgtgctcttccgatct**GCTAC**ACTACTACTCTGTATGGTTGGT |
| S_Rbd_V2_Rev_well-BC_2 | gtgactggagttcagacgtgtgctcttccgatct**TGACG**ACTACTACTCTGTATGGTTGGT |
| S_Rbd_V2_Rev_well-BC_3 | gtgactggagttcagacgtgtgctcttccgatct**ATCGT**ACTACTACTCTGTATGGTTGGT |
| S_Rbd_V2_Rev_well-BC_4 | gtgactggagttcagacgtgtgctcttccgatct**CAGTA**ACTACTACTCTGTATGGTTGGT |
| S_Rbd_V2_Rev_well-BC_5 | gtgactggagttcagacgtgtgctcttccgatct**GTCAC**ACTACTACTCTGTATGGTTGGT |
| S_Rbd_V2_Rev_well-BC_6 | gtgactggagttcagacgtgtgctcttccgatct**TAGCG**ACTACTACTCTGTATGGTTGGT |
| S_Rbd_V2_Rev_well-BC_7 | gtgactggagttcagacgtgtgctcttccgatct**ACTGT**ACTACTACTCTGTATGGTTGGT |
| S_Rbd_V2_Rev_well-BC_8 | gtgactggagttcagacgtgtgctcttccgatct**CGATA**ACTACTACTCTGTATGGTTGGT |
| S_Rbd_V2_Rev_well-BC_9 | gtgactggagttcagacgtgtgctcttccgatct**GTTGC**ACTACTACTCTGTATGGTTGGT |
| S_Rbd_V2_Rev_well-BC_10 | gtgactggagttcagacgtgtgctcttccgatct**TAATG**ACTACTACTCTGTATGGTTGGT |
| S_Rbd_V2_Rev_well-BC_11 | gtgactggagttcagacgtgtgctcttccgatct**ACGAT**ACTACTACTCTGTATGGTTGGT |
| S_Rbd_V2_Rev_well-BC_12 | gtgactggagttcagacgtgtgctcttccgatct**CGTCA**ACTACTACTCTGTATGGTTGGT |
| S_Rbd_V2_Rev_well-BC_13 | gtgactggagttcagacgtgtgctcttccgatct**GTAGC**ACTACTACTCTGTATGGTTGGT |
| S_Rbd_V2_Rev_well-BC_14 | gtgactggagttcagacgtgtgctcttccgatct**TACTG**ACTACTACTCTGTATGGTTGGT |
| S_Rbd_V2_Rev_well-BC_15 | gtgactggagttcagacgtgtgctcttccgatct**AGTAC**ACTACTACTCTGTATGGTTGGT |
| S_Rbd_V2_Rev_well-BC_16 | gtgactggagttcagacgtgtgctcttccgatct**CTACG**ACTACTACTCTGTATGGTTGGT |
| S_Rbd_V2_Rev_well-BC_17 | gtgactggagttcagacgtgtgctcttccgatct**GACGT**ACTACTACTCTGTATGGTTGGT |
| S_Rbd_V2_Rev_well-BC_18 | gtgactggagttcagacgtgtgctcttccgatct**TCGTA**ACTACTACTCTGTATGGTTGGT |
| S_Rbd_V2_Rev_well-BC_19 | gtgactggagttcagacgtgtgctcttccgatct**AGCAC**ACTACTACTCTGTATGGTTGGT |
| S_Rbd_V2_Rev_well-BC_20 | gtgactggagttcagacgtgtgctcttccgatct**CTGCG**ACTACTACTCTGTATGGTTGGT |
| S_Rbd_V2_Rev_well-BC_21 | gtgactggagttcagacgtgtgctcttccgatct**GATTG**ACTACTACTCTGTATGGTTGGT |
| S_Rbd_V2_Rev_well-BC_22 | gtgactggagttcagacgtgtgctcttccgatct**TCAAT**ACTACTACTCTGTATGGTTGGT |
| S_Rbd_V2_Rev_well-BC_23 | gtgactggagttcagacgtgtgctcttccgatct**AGCCA**ACTACTACTCTGTATGGTTGGT |
| S_Rbd_V2_Rev_well-BC_24 | gtgactggagttcagacgtgtgctcttccgatct**CTGGC**ACTACTACTCTGTATGGTTGGT |

**Supplementary Table 4: *S-Rbm*-well-BC primer list.**

| Variant of Concern | Woldwide Emergence (Locatio | Worldwide Emergence (Date) | First Case Detected by C19-SPAR-Seq |
|---|---|---|---|
| Alpha (B1.1.7) | United Kingdom | September 2020 | December 2020 |
| Beta (B.1.351) | South Africa | May 2020 | December 2020 |
| Gamma (P.1) | Brazil | November 2020 | January 2021 |
| Delta (B.1.617.2) | India | October 2020 | June 2021 |
| Omicron (B.1.1.529) | South Africa | November 2021 | November 2021 |
| Omicron (BA.2) | South Africa | November 2021 | December 2021 |
| Omicron (BA.3) | South Africa | November 2021 | December 2021 |
| Omicron (BA.4) | South Africa | January 2022 | May 2022 |
| Omicron (BA.5) | South Africa | February 2022 | May 2022 |
| Omicron (XBB) | India | August 2022 | November 2022 |
| Omicron (XBB. 1.5) | United States | October 2022 | December 2022 |

**Supplementary Table 5. Date of first detection of global variants of concern within Toronto.**

| Date | # Fig 3 | status | Milestone | Web link |
|---|---|---|---|---|
| January 23th, 2021 | 1 | Closing | Stay-at-home order for all Ontario, eg: Province under a provincewide Stay-at-Home order, requiring everyone to remain at home except for essential purposes, such as going to the grocery store or pharmacy, accessing health care services (including getting vaccinated), for outdoor exercisewith your household in your home community, or for work that cannot be done remotely | https://news.ontario.ca/en/release/60261/ontario-extending-stay-at-home-order-across-most-of-the-province-to-save-lives |
| February 8th, 2021 | 2 | Opening | Return to the classroom in-person learning for regions other than Peel, Toronto, York | https://toronto.citynews.ca/2021/03/11/timeline-a-year-of-pandemic-life/ |
| February 16th, 2021 | 3 | Opening | Stay-at-home order lifted; return to classroom in-person learning for Peel, Toronto, York | https://news.ontario.ca/en/release/60228/enhanced-safety-measures-in-place-as-in-person-learning-resumes-across-ontario |
| February 22nd, 2021 | 4 | Closing | Back to stay-at-home order | https://news.ontario.ca/en/release/60261/ontario-extending-stay-at-home-order-across-most-of-the-province-to-save-lives |
| March 8th, 2021 | 5 | Opening | Stay-at-home order lifted | https://news.ontario.ca/en/release/60261/ontario-extending-stay-at-home-order-across-most-of-the-province-to-save-lives |
| April 3rd, 2021 | 6 | Closing | Return to stay at home | https://toronto.citynews.ca/2021/03/08/covid-19-grey-zone-toronto-peel/ |
| April 19 2021 | 7 | Closing | Return to remote learning | https://news.ontario.ca/en/release/61106/ontario-moves-schools-to-remote-learning-following-spring-break |
| June 1st, 2021 | 8 | | Start of daily SPAR-seq testing | https://globalnews.ca/news/7743332/ontario-covid-stay-at-home-order/ |
| June 2 2021 | 9 | Closing | Ontario continues remote learning until end of school year | https://news.ontario.ca/en/release/1000251/remote-learning-to-continue-across-ontario-for-the-remainder-of-school-year |
| June 11th, 2021 | 10 | Opening | Reopening phase 1 including loosening of restrictions to outdoor areas/gatherings ("Step 1 of the Roadmap To Reopen plan") (https://news.ontario.ca/en/backgrounder/1000159/roadmap-to-reopen) | https://news.ontario.ca/en/release/1000279/ontario-to-move-to-step-one-of-roadmap-to-reopen-on-june-11 |
| June 30th, 2021 | 11 | Opening | Reopening phase 2 including loosening of restrictions in some indoor and further outdoor areas ("Step 2 of the Roadmap To Reopen plan") (https://news.ontario.ca/en/backgrounder/1000159/roadmap-to-reopen) | https://news.ontario.ca/en/release/1000399/ontario-moving-to-step-two-of-roadmap-to-reopen-on-june-30 |
| July 16 2021 | 12 | Opening | Reopening phase 3 including futher loosing of resrictions in indoor areas ("Step 3 of the Roadmap To Reopen plan") (https://news.ontario.ca/en/backgrounder/1000159/roadmap-to-reopen) | https://news.ontario.ca/en/release/1000501/ontario-moving-to-step-three-of-roadmap-to-reopen-on-july-16 |
| Sep 8 2021 | 13 | Opening | Return to in person learning | https://www.ontario.ca/document/covid-19-health-safety-and-operational-guidance-schools-2021-2022 |
| Oct 25 2021 | 14 | Opening | Ontario will lift capacity limits, including physical distancing requirements, in the vast majority of settings where proof | https://news.ontario.ca/en/release/1001027/ontario-releases-plan-to- |
| November 30th, 2021 | 15 | Closing | Border measures: Entry prohibitions for travellers coming from 13 African countries as response to first reported | https://www.ctvnews.ca/health/coronavirus/canada-bans-travellers- |
| Dec 19 2021 | 16 | Closing | Ontario returns to restrictions on indoor settings as response to increasing cases | https://toronto.ctvnews.ca/ontario-slashing-gathering-sizes-reducing- |
| December 31th, 2021 | 17 | | End of systematic COVID19 screening for the public | https://news.ontario.ca/en/backgrounder/1001387/updated-eligibility- |
| January 5 2022 | 18 | Closing | Return to step 2 of "Roadmap to Reopen Plan", schools closed | https://news.ontario.ca/en/release/1001394/ontario-temporarily- |

**Supplementary Table 6. List of dates and descriptions of non-pharmaceutical interventions and other relevant events in the GTA from January 2021 to January 2022.**

| Variant | Input Samples (total 1110) | Samples producing an S-Rbm sequence | S-Rbm Output Sequences (total count, not unique) | S-Rbm Unique Output Sequences | Samples producing an S-Pbs sequence | S-Pbs Output Sequences (total count, not unique) | S-Pbs Unique Output Sequences |
|---|---|---|---|---|---|---|---|
| WT | 301 | **145 (48.17%)** | **225** | **13** | **1 (0.33%)** | **1** | **1** |
| Alpha | 75 | **38 (50.67%)** | **63** | **5** | **4 (5.33%)** | **4** | **1** |
| Delta | 193 | **12 (6.23%)** | **15** | **6** | **6 (3.11%)** | **7** | **4** |
| Omicron B.1.1.529 | 541 | **33 (6.1%)** | **38** | **5** | **10 (1.85%)** | **10** | **2** |

**Supplementary Table 7. Summary of samples from different VOCs used for pQS analysis and the sequences outputted by the analysis.**
